# Global risk of selection and spread of *Plasmodium falciparum* histidine-rich protein 2 and 3 gene deletions

**DOI:** 10.1101/2023.10.21.23297352

**Authors:** Oliver J. Watson, Thu Nguyen-Anh Tran, Robert J Zupko, Tasmin Symons, Rebecca Thomson, Theodoor Visser, Susan Rumisha, Paulina A Dzianach, Nicholas Hathaway, Isaac Kim, Jonathan J. Juliano, Jeffrey A. Bailey, Hannah Slater, Lucy Okell, Peter Gething, Azra Ghani, Maciej F Boni, Jonathan B. Parr, Jane Cunningham

## Abstract

In the thirteen years since the first report of *pfhrp2*-deleted parasites in 2010, the World Health Organization (WHO) has found that 40 of 47 countries surveyed worldwide have reported *pfhrp2/3* gene deletions. Due to a high prevalence of *pfhrp2/3* deletions causing false-negative HRP2 RDTs, in the last five years, Eritrea, Djibouti and Ethiopia have switched or started switching to using alternative RDTs, that target pan-specific-pLDH or *P. falciparum* specific-pLDH alone of in combination with HRP2. However, manufacturing of alternative RDTs has not been brought to scale and there are no WHO prequalified combination tests that use Pf-pLDH instead of HRP2 for *P. falciparum* detection. For these reasons, the continued spread of *pfhrp2/3* deletions represents a growing public health crisis that threatens efforts to control and eliminate *P. falciparum* malaria. National malaria control programmes, their implementing partners and test developers desperately seek *pfhrp2/3* deletion data that can inform their immediate and future resource allocation. In response, we use a mathematical modelling approach to evaluate the global risk posed by *pfhrp2/3* deletions and explore scenarios for how deletions will continue to spread in Africa. We incorporate current best estimates of the prevalence of *pfhrp2/3* deletions and conduct a literature review to estimate model parameters known to impact the selection of *pfhrp2/3* deletions for each malaria endemic country. We identify 20 countries worldwide to prioritise for surveillance and future deployment of alternative RDT, based on quickly selecting for *pfhrp2/3* deletions once established. In scenarios designed to explore the continued spread of deletions in Africa, we identify 10 high threat countries that are most at risk of deletions both spreading to and subsequently being rapidly selected for. If HRP2-based RDTs continue to be relied on for malaria case management, we predict that the major route for *pfhrp2* deletions to spread is south out from the current hotspot in the Horn of Africa, moving through East Africa over the next 20 years. We explore the variation in modelled timelines through an extensive parameter sensitivity analysis and despite wide uncertainties, we identify three countries that have not yet switched RDTs (Senegal, Zambia and Kenya) that are robustly identified as high risk for *pfhrp2/3* deletions. These results provide a refined and updated prediction model for the emergence of *pfhrp2/3* deletions in an effort to help guide *pfhrp2/3* policy and prioritise future surveillance efforts and innovation.

## Introduction

The expanded use of malaria rapid diagnostic tests (RDTs) in the last twenty years has been central to global malaria control efforts to test, treat and track all malaria infections, with 262 million RDTs distributed in 2021 by national malaria programmes (NMPs) and 413 million sold by WHO prequalified manufacturers (*1*). The RDTs commonly deployed for diagnosis of falciparum malaria detect *Plasmodium falciparum* histidine-rich protein 2 (PfHRP2) and its paralog *P. falciparum* histidine-rich protein 3 (PfHRP3). However, progress against malaria is now threatened by an increase in *pfhrp2/3* gene deletions resulting in false-negative RDT results. In 2014, a review was conducted that called for a harmonised approach to investigate and report *pfhrp2/3* gene deletions (*2*). As of 2023, the World Health Organization (WHO) Malaria Threat Maps includes reports of *pfhrp2/3* deletions in 40 of 47 countries surveyed worldwide (*3*), and report of pfhrp2 deletions causing false-negative rates have been as high as 80% in the worst affected settings (*4*). Once detected, there are concerns that *pfhrp2/3* deletions may be rapidly selected for, as demonstrated by observations in Eritrea and Ethiopia (*4*, *5*). There are alternative, non-HRP2 based RDTs that target alternative antigens such as *Plasmodium* lactate dehydrogenase (pLDH). Pan-specific-pLDH RDTs, however, have not been brought to scale because of their lower sensitivity compared to HRP2 and for countries that need to both detect and distinguish between *P. falciparum* and *P. vivax*, there are no WHO prequalified combination tests that use Pf-pLDH instead of or in addition to HRP2 for *P. falciparum* detection. This has posed particular challenges as *pfhrp2/3* deletions have emerged and become dominant in several countries that require this type of combination product e.g. Eritrea, Ethiopia, Djibouti, Peru, Brazil. Most other countries continue to rely on PfHRP2-based RDTs as their primary malaria diagnostic tool, so emergence and spread of *pfhrp2/3-*deleted strains represents a growing public health crisis and poses a significant obstacle to the control and eradication of *P. falciparum*.

Accurate maps of *pfhrp2/3-*deleted strains and their impact on HRP2 RDT results are needed to understand the current risk to malaria control but also to parameterise the risk of future spread. Multiple molecular surveys have been undertaken to characterise the current spread and estimated prevalence of parasites with *pfhrp2/3* deletions (genotype frequency of *pfhrp2/3* deletions). However, accurately estimating both the true frequency of *pfhrp2/3* deletions, their impact on HRP2-RDT results and the risk they pose to malaria control is challenging. One challenge is the need to harmonize estimates of *pfhrp2/3* deletions across studies with different sampling and laboratory testing schemes, which prompted the WHO to publish methodological guidance and protocols in 2018 for studying *pfhrp2/3* deletions (*6*). However, a review of published surveys (*7*) concluded that unrepresentative surveys and inconsistent study design has impaired efforts to evaluate the risk of *P. falciparum* malaria cases being misdiagnosed due to *pfhrp2/3* deletions. Additionally, more recent surveys with newer laboratory techniques for detecting *pfhrp2/3* deletions have detected lower frequencies of *pfhrp2/3* deletions (*8*) than previous surveys (*9*). Secondly, evidence suggested that there are differences in the phenotypes associated with deleted parasites between geographical regions. For example, in the Democratic Republic of Congo (DRC), a high level of deletions (6.4%, 95% confidence interval 5.1-8.0%) was found when using asymptomatic samples from Demographic and Health Surveys (DHS) (*10*). However, a subsequent study in symptomatic patients using improved laboratory methods in the same regions found no symptomatic malaria cases with *pfhrp2* deletions (*11*). In contrast, Eritrea (*4*, *12*) and Djibouti (*13*, *14*) are affected by a high frequency of *pfhrp2/3-*deleted parasites that cause symptomatic and clinically relevant infections. Further, in Peru, deleted parasites emerged in settings that have never relied upon HRP2-based RDTs for diagnosis, prompting speculation that deletions offer an as yet undefined selective advantage in this context beyond evasion of diagnosis (*15*). These distinct phenotypes imply different immediate risks to malaria control and suggest that different evolutionary pressures are driving heterogenous spread of *pfhrp2/3* deletions across regions (*16*).

In 2017, an individual-based mathematical model of malaria transmission characterising the drivers of selection for *pfhrp2* deletions was developed (*17*). However, there was insufficient data to comprehensively account for the impact of *pfhrp2* gene deletions on parasite fitness and the different mechanisms of selection driving the distinct spread between *pfhrp2* and *pfhrp3* deletions. Additionally, limited data were available to parameterise the proportion of malaria cases diagnosed by microscopy, the level of adherence to RDT-based treatment, the cross-reactivity of HRP3 epitopes to yield a positive HRP2-RDT, and the incidence of non-malarial febrile illness (NMF) - all factors expected to impact the selective advantage of *pfhrp2* deletions. However, new studies and data provide improved insight into these processes. For example, HRP3 cross-reactivity has been shown to be higher than previously thought, with HRP3 cross-reactivity on HRP2-based RDTs sufficient to mask the effects of *pfhrp2* deletions in high parasite density in-vitro cultures (*18*). However, cross reactivity will differ between brands depending on the target epitopes of the antibodies bound to the test strips (*19*) and target field data from malaria symptomatic patients in Ethiopia showed different performance with 46% (12/26) of *pfhrp2−/3+* samples yielding a positive HRP2-based RDT (*5*). With regards to the evolutionary mechanism driving *pfhrp2/3* selection, population genetic analyses conducted in Ethiopia concluded that *pfhrp3* deletion has arisen independently multiple times, whereas *pfhrp2* deletion likely arose more recently due to the strong positive selection owing to HRP2-RDT-based test-and-treat policy (*5*). Understanding how strongly *pfhrp2* deletion is linked to *pfhrp3* deletion is critical - if these two deletions co-occur more than would be expected by chance (analogous to positive linkage disequilibrium [LD] but between genes on different chromosomes), the benefits for RDT performance conferred by HRP3 cross reactivity will be negated. Lastly, *in vitro* competition assays of asexual parasite fitness suggest that up to a 90% relative fitness (a 10% loss in replicative rate) may be associated with *pfhrp2* deletions (*20*), although no *in vivo* nor feeding assay studies have been conducted to assess fitness costs throughout the parasite life cycle.

In this study, we incorporate recent advances in our understanding of *pfhrp2/3* deletions and new data relevant to their spread to provide a global assessment of the risk posed by *pfhrp2/3*-deleted parasite strains. We evaluate the susceptibility of each region to spread after deleted parasites are established and predict the spread of *pfhrp2/3* deletions globally with a focus on sub-Saharan Africa (SSA) based on best estimates of the prevalence of *pfhrp2* deletions. These risk maps can be used to guide ongoing surveillance efforts, future deployment of alternative RDTs, and research to improve our understanding of the biology and threat of *pfhrp2/3* deletions.

## Methods

### *P. falciparum* transmission model

In this study, we employed a previously developed individual-based mathematical model of *P. falciparum* malaria transmission to simulate the selection of *pfhrp2* deletions (*17*). The model monitors the transmission of *pfhrp2*-deleted parasites and wild-type parasites (i.e. *pfhrp2* positive) between human and mosquito hosts. We describe the model in brief here before detailing further considerations related to *pfhrp3* dynamics and the data sources used to parameterise the model for simulating *pfhrp2* deletions globally.

Individuals are born with maternally acquired immunity that decays within the first six months, rendering them susceptible to infection from infectious mosquito bites. Exposure depends on the entomological inoculation rate (EIR), which is location-specific. Upon infection, individuals acquire either a *pfhrp2*-deleted parasite or a wild-type parasite. This is determined by the genotype frequency of *pfhrp2-*deleted parasites in humans 30 days prior, which accounts for the lags between human exposure, parasite gametocytogenesis and sporozoite development in mosquitoes. Following a short latent period, infected individuals either develop clinical symptomatic disease (probability determined by their level of blood-stage immunity, with immunity increasing with age and exposure) or progress as an asymptomatic infection. Symptomatic individuals may seek treatment, and they are assumed to be successfully treated unless they are infected with only *pfhrp2* deleted parasites and the decision to treat is determined only by a positive HRP2-based RDT. All other possible outcomes from an individual seeking treatment (nonadherence to negative RDT outcome, positive HRP2-based RDT due to cross reactivity with HRP3 epitopes, microscopy or alternative RDT (not exclusively reliant on HRP2), used for diagnosis or the individual is treated without being tested) result in the individual being successfully treated. Once treated, individuals undergo a prophylactic period before returning to susceptibility. Asymptomatically infected individuals recover more slowly, with detectability influenced by immunity levels. Super-infection is incorporated, with asymptomatically infected individuals exposed at the same rate as susceptible individuals. Acquired strains from previous infection are naturally cleared after a period similar to the duration of an asymptomatic infection that has not been extended due to super-infection. All infected states are infectious to mosquitoes, with infectivity dependent on detectability (serving as a surrogate for asexual parasite density). Mosquitoes become infected at a rate dependent on human population infectivity and become infectious after approximately 10 days, reflecting the extrinsic incubation period. The model has been parameterized by fitting it to data on the relationship between EIR, parasite prevalence, clinical disease incidence, and severe disease incidence. The model has also been shown to accurately capture the selection and relationship between *pfhrp2* deletion frequency and transmission intensity in the DRC (*17*). Full mathematical details are available in Watson et al. (*17*).

#### *Pfhrp3* dynamics

In a previous modelling analysis, we assumed a fixed probability of 25% that an individual infected with parasites with only *pfhrp2* deleted (i.e. *pfhrp3* present) would test positive by HRP2-based RDTs due to cross reactivity with HRP3 epitopes. To more accurately capture the role of *pfhrp3*, we conducted a scoping review of RDT performance on *pfhrp2-/pfhrp3+* clinical infections to estimate the probability that a positive RDT would occur if *pfhrp3* is present. Secondly, we note that *pfhrp3* deletions are frequently found at higher frequencies than *pfhrp2* deletions despite *pfhrp2* deletions providing a greater advantage than *pfhrp3* deletions with regards to the ability to evade diagnosis by HRP2-based RDTs (*21*). This observation reflects the mechanistic (*22*) and soft selective processes that are hypothesised to result in the emergence of *pfhrp3* deletions (*5*). This observation is in contrast to the strong selective sweeps associated with *pfhrp2* deletions due to RDT-based test-and-treatment that cause *pfhrp2* deletions to be selected on both genetic backgrounds, but more strongly on a *pfhrp3-deleted* background (*5*). Consequently, we continue to only explicitly model *pfhrp2* deletions in our model and estimate the probability that a *pfhrp2* deleted parasite has an intact *pfhrp3* gene. If *pfhrp3* is intact, the probability that an individual will yield a positive HRP2-based RDT is determined by the probability of HRP3 cross reacting, which is estimated later as part of a model fitting exercise. In effect, we model the probability that an individual whose parasites have only *pfhrp2* deletions would have circulating HRP3 due to intact *pfhrp3* and that these yield a positive HRP2-based RDT due to cross-reactivity with HRP3 epitopes.

To estimate the association or linkage disequilibrium (between genes on different chromosomes) between *pfhrp2* and *pfhrp3* deletions, we used the WHO Malaria Threat Map data to calculate, per study, the total number of *pfhrp2-/pfhrp3-*, *pfhrp2-/pfhrp3+*, *pfhrp2+/pfhrp3-* and *pfhrp2+/pfhrp3*+ samples. From the resultant two-by-two table, we calculate the normalised coefficient of linkage disequilibrium, *D*’, given by:

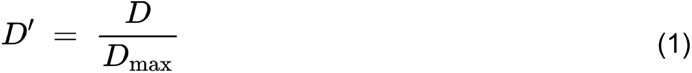

Where *D* is the coefficient of linkage disequilibrium, and *D*_*max*_ is the theoretical maximum difference between the observed and expected haplotype frequencies, given by:

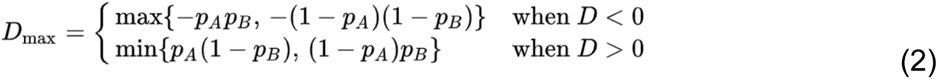

To estimate the likelihood that *pfhrp2* deletions arise without *pfhrp3* deletions, we calculated the proportion of all *pfhrp2* deleted infections without *pfhrp3* deletions. For each continent, we fit a Beta Binomial distribution (to account for overdispersion across studies) to the calculated study proportions, with the estimated mean used to represent the probability that *pfhrp2* deletions would arise without *pfhrp3* deletions.

### Model parameters for modelling the selection of *pfhrp2* globally

Based on previous modelling efforts, we identified a list of risk factors that impact the speed of selection of *pfhrp2* deletions (**Table 1**). We conducted an extensive literature and database review to source estimates for each of the risk factors at the first administrative unit (or national level if not available subnationally) for all countries with stable malaria transmission. A full description of the review, including search terms, inclusion criteria and how the resultant data source used was incorporated is available in the Supplementary Methods.

**Table 1:** Model parameters that impact the speed of selection for *pfhrp2/3* deletions and data sources used to estimate these parameters. All parameters were sourced at the national level, except for malaria prevalence and treatment-seeking rates, which were sourced at the first administrative unit. See Supplementary Methods for full description.

| Drivers of *pfhrp2/3* selection | Impact on speed of selection for *pfhrp2/3* deletions | Data Sources Used |
| --- | --- | --- |
| Malaria Prevalence | Lower malaria prevalence will increase selection pressure by increasing the probability that individuals are only infected with *pfhrp2/3* deleted parasites and thus more likely to not be treated. Additionally, lower malaria prevalence will increase the probability that an infected individual will develop a symptomatic infection (due to lower immunity at lower transmission intensities), which in turn influences the infected individual's likelihood to seek treatment. | Malaria Atlas Project maps of slide positivity ages 2-10 (23). |
| Microscopy-based diagnosis | The use of microscopy for malaria diagnosis will decrease selection pressure by negating the selective advantage conferred by *pfhrp2/3* deletions. | WHO World Malaria Report "proportion of cases confirmed by diagnostic" table, with missing data imputed using all other collected model parameters. |
| Treatment-seeking rate for fever | Increased treatment-seeking will increase the rate at which the selective advantage conferred by *pfhrp2/3* is able to be realised by evading diagnosis and treatment. | Commodities Forecast Dashboard by the Malaria Atlas Project (25), which uses Demographic and Health Surveys (DHS), Malaria Indicator Surveys (MIS), Multiple Indicator Cluster Surveys (MICS) and AIDS Indicator Surveys (AIS) in generalized additive mixed model (GAMM) to predict treatment seeking patterns over time. |
| Proportion of treatment-seeking for fever in the private sector. | Low use of malaria rapid diagnostic tests has been shown to exist in the private market in a number of locations (24). If the use of RDTs is lower in the private market than in the public sector then selection pressure will decrease with an increasingly large private drug market. | DHS/MIS Surveys used in GAMM for estimating treatment seeking from any (medical) source and for estimating treatment seeking in the public sector. |
| Proportion of individuals seeking care who receive diagnostic test | Low use of any diagnostic test for guiding treatment decisions will reduce selection pressure for *pfhrp2/3* deletions. | DHS data (surveys in Africa asking if care-seeking febrile children received a finger/heel prick). |
| Nonadherence to RDT outcomes | Nonadherence to RDT outcomes (treating RDT negative individuals) will decrease selection pressure by negating the selective advantage conferred by *pfhrp2/3* deletions. | Commodities Forecast Dashboard by the Malaria Atlas Project (25), which uses a statistical model of the probability of care-seeking fevers receiving any antimalarial informed by DHS and MIS data. |
| RDT brands | The use of non-HRP2-based RDTs will negate the selective advantage conferred by *pfhrp2/3* deletions. | Global Fund Price and Quality Reporting and President's Malaria Initiative data on volumes of RDT test types and brands used. |
| Cross-reactivity of PfHRP3 epitopes | Increasing cross-reactivity between PfHRP3 epitopes and PfHRP2-based RDTs will decrease selection pressure for *pfhrp2* deletions. | Estimate based upon WHO Malaria Threat Maps data and studies reporting performance of HRP2-based RDTs on *pfhrp2*-/ *pfhrp3*+ ((5), (14), (26)). |
| Fitness costs associated with *pfhrp2/3* gene deletions. | Fitness costs associated with *pfhrp2/3* gene deletions will reduce the transmissibility of gene deleted parasites. | Parameterised via model fitting to Eritrean and Ethiopian *pfhrp2/3* deletion data, with priors from *in vitro* competition assay data (20). |

#### Refining estimates of fitness costs associated with *pfhrp2* deletions

One notable uncertainty for modelling *pfhrp2-*deleted parasites is whether deleted parasites suffer a fitness cost and how that fitness cost impacts the probability of deleted parasites being onwardly transmitted. Asexual fitness costs have been measured by conducting pairwise competition experiments *in vitro*, suggesting a fitness cost of 8.7% (relative fitness of 91.3%) for *pfhrp2* deleted parasite strains and 11.3% (relative fitness of 88.7%) for strains with both *pfhrp2* and *pfhrp3* deletions (*20*). These fitness costs were estimated by comparing the growth of *pfhrp2* and/or *pfhrp3* knocked-out strains against a common competitor strain. Consequently, the inferred fitness costs reflect the impact on asexual parasite growth in mixed infections. However, it is unknown whether these measured fitness costs translate to a reduction in onward infection (how we model parasite fitness costs). Additionally, previous feeding assay studies have highlighted the importance of measuring both the fitness of asexual and sexual stages to fully characterise the impact on population level trends (*27*).

To estimate the fitness costs associated with *pfhrp2* deletions in our model, we used our transmission model to model the selection of *pfhrp2* deletions in Eritrea and Ethiopia at each first administrative unit. We chose Eritrea and Ethiopia for this parameter estimation exercise as both countries contain multiple surveys and represent known “hot spots” of *pfhrp2/3* deletions in Africa that have also been shown to cause symptomatic infection. In addition, the surveys include data on *pfhrp3* deletions, which allow for the probability that *pfhrp2* deletions occur with *pfhrp3* deletions to be estimated for each location (revealing that *pfhrp2* deletions rarely were observed without *pfhrp3* deletions). We chose not to include Djibouti in this exercise due to uncertainty in recent malaria prevalence and the impact of outbreaks due to the emerging spread of *Anopheles stephensii* (*28*).

We statistically compared the modelled frequency of *pfhrp2* deletions against representative *pfhrp2* surveys from the WHO Malaria Threat Maps to jointly infer parameter values for both the comparative fitness costs and the cross reactivity of HRP3 epitopes. We used a Bayesian approach, with a flat prior for the fitness cost, with bounds centred on the fitness cost estimated in the *in vitro* fitness study (*20*) (relative fitness parameter bound between 0.8 - 0.99) and a Beta distribution (alpha = 13, beta = 15) for the probability of HRP3 cross reacting informed. This prior was informed by studies of the performance of HRP2-based RDTs on *pfhrp2-/pfhrp3+* samples in Ethiopia, which observed 46.2% (12/26) of samples yielding a positive RDT (*5*). While other studies in Djibouti (*14*) and Uganda (*26*) reported lower cross reactivity (0/5 and 1/10 samples cross reacting respectively), we chose a prior based on the Ethiopian study given the location of the *pfhrp2* surveys we are fitting to in Eritrea and Ethiopia and because no data was available in Eritrea due to previous studies either only observing *pfhrp2-/pfhrp3-* samples or not testing *pfhrp2-/pfhrp3+* samples with RDT. Log-likelihoods were calculated for each study by assuming the proportion of *pfhrp2* deletions was described by a Binomial distribution, with the number of samples genotyped in each study used as the number of trials. Median estimates and 95% credible intervals for each parameter were obtained from 1000 draws from the posterior parameter space (see Supplementary Methods for full model fit details).

### *Pfhrp2* deletion risk scores

In our previous analysis, we created risk scores of “HRP2 Concern”. To create these scores, we simulated trends in the prevalence of *pfhrp2*-deleted mutants across SSA. These simulations included estimates of the mean microscopy-based, *Plasmodium falciparum* prevalence in 2–10-year-olds (*Pf*PR_2-10_) in 2010 by first administrative unit, and estimates of the proportion of cases seeking treatment from previously modelled estimates using the DHS and the Malaria Indicator Cluster Surveys (*29*). The time taken for the proportion of infections with all strains *pfhrp2-*deleted to reach 20% was recorded and classified to map areas of HRP2 concern under four qualitative classifications. This approach, however, relied on a different metric (namely the proportion of infections with all strains *pfhrp2-*deleted) to the 5% false-negative RDTs due to pfhrp2 deletions metric subsequently adopted by the WHO for deciding when to switch RDTs (*30*). This metric is based on the proportion of clinically relevant infections that would be misdiagnosed due to *pfhrp2/3* gene deletions. To address this discrepancy, we produce maps of two new risk scores - the “Innate Risk Score” and the “Prospective Risk Score” - based on the proportion of clinically relevant infections that would be misdiagnosed due to *pfhrp2/3* gene deletions. To create these new maps, we use updated estimates from 2020 for the parameters described in **Table 1** and assume that these estimates remain constant going forwards, i.e. malaria transmission intensity, treatment-seeking data, and RDT usage data remain the same as estimated in 2020.

#### Innate Risk Score

The first risk score, the “Innate Risk Score”, is the innate potential for *pfhrp2* deletions to spread once established in a region based solely on the region’s malaria transmission intensity, treatment-seeking data, and adherence to diagnostic test outcome. Informed by the current 5% WHO threshold, we define the Innate Risk Score as the time taken for the percentage of clinical cases to be misdiagnosed by PfHRP2-based RDTs to increase from 1% (previously shown to be a suitable threshold for defining establishment of *P. falciparum* genetic traits under positive selection (*31*)) to 5%. We then use a similar approach as in Watson et al. (*17*) to categorise each region’s Innate Risk Score. Here, a region’s risk is classified as High, Moderate or Slight, defined as reaching the 5% threshold within 6, 12 and 20 years, respectively, or marginal risk if 5% is not reached within 20 years. Importantly, we do not incorporate data on the current types of RDT used in that country (these are used in the “Prospective Risk Score”). Consequently, the Innate Risk Score reflects the risk that deletions would spread in a region if all types of RDT used were HRP2-based RDTs. While the majority of countries continue to use only HRP2-based RDTs, a number of countries in SSA have switched to non-HRP2-based RDTs, Eritrea, Djibouti and partially Ethiopia In these countries, the Innate Risk Score thus conveys the risk that is still posed if those countries reverted back to only HRP2-based RDTs.

To estimate the Innate Risk Score for each administrative level 1 region, we first estimate the selection coefficient (the annual % change in logit genotype frequency (*32*)) for clinical cases to be misdiagnosed by PfHRP2-based RDTs. We estimate selection coefficients using the following approach, which is expanded fully in the Supplementary Methods. We first created 8,748 unique parameter sets that equally span the range observed globally for each model parameter detailed in **Table 1**. For all parameter combinations, five stochastic realisations of 100,000 individuals were simulated for 40 years to reach equilibrium first before simulating the selection of *pfhrp2* deletions over the following 20 years, with a starting frequency of *pfhrp2* deletions equal to 6%. 6% was chosen based on recommendations made by a previous modelling study (*32*), which recommends selecting an allele frequency as low as possible to reflect the condition under which most selection occurs but also high enough to reduce stochastic noise in allele spread and allow for more accurate estimation of selection coefficients from modelling outputs. For each simulation we calculate the selection coefficient (*32*) associated with the increase in clinical cases misdiagnosed by HRP2-based RDTs due to *pfhrp2/3* deletions (**Figure 1**) (*33*, *34*).

**Figure 1.**
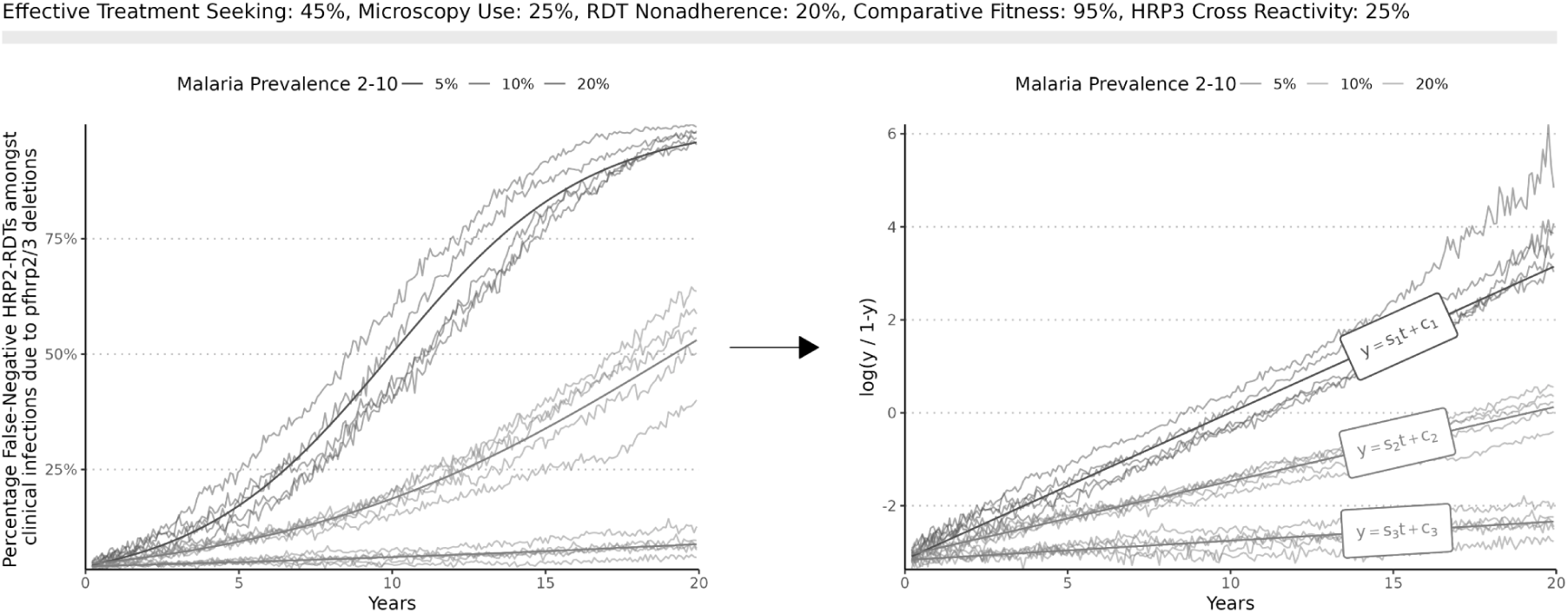
Conversion from model simulations to selection coefficients. For a given parameter set (effective treatment-seeking: 45%, microscopy use: 25%, RDT nonadherence: 20%, comparative fitness: 95%, HRP3 cross reactivity: 25%), the simulated percentage of false-negative HRP2-based RDTs amongst clinical infections due to *pfhrp2/3* deletions (y) is converted to log odds (y/1-y), with the gradient calculated to estimate the selection coefficient.

We next train an ensemble machine learning model to predict selection coefficients based on model simulation parameters (malaria prevalence, effective treatment-seeking as a result of the treatment cascade, adherence to RDT outcomes for decision to treat, microscopy use for diagnosis, comparative fitness costs of *pfhrp2* deletions, and probability of testing positive by HRP2-based RDT despite being *pfhrp2* deleted due to intact *pfhrp3* and cross-reactivity with HRP3 epitopes). This approach provides a statistical model that replicates the underlying transmission model behaviour that can be subsequently generalised to any transmission setting. From these models, we predict how quickly the 5% threshold will be reached once *pfhrp2* deletions are established in a region (defined as 1% frequency based on previous antimalarial resistance modelling exercises (*31*)). Uncertainty in selection coefficients due to stochastic variation in model simulations was also estimated using a similar statistical modelling framework. We trained a statistical model to predict the variation in selection coefficients observed across stochastic realisations for a particular parameter set.

#### Prospective Risk Score

The Innate Risk Score, while capturing the underlying selection dynamics, does not incorporate data on the current distribution of *pfhrp2/3* deletions in Africa. The second risk score, which we call the “Prospective Risk Score”, is calculated from a prospective modelling approach designed to explore different scenarios for how *pfhrp2* deletions may continue to spread in Africa based on current estimates of the prevalence of *pfhrp2* deletions from the WHO Malaria Threat Maps. While there are considerable uncertainties in the prevalence of gene deletions across Africa (*7*) and identifying the true denominator in reported surveys is challenging (*21*), these estimates represent our best understanding of the current genotype frequency of *pfhrp2* deletions in Africa. In countries without molecular surveillance data, we assume the current frequency of *pfhrp2* deletions is 0%.

Given the difficulty in estimating the rate at which malaria parasites under selection spread geographically (*35*), we use a simple model of parasite movement to describe how *pfhrp2/3* deletions spread between administrative regions. To simulate the spread between regions, we make the simplifying assumption that *pfhrp2* deletions are exported from an admin level 1 region once *pfhrp2* deletions are found in 25% of clinical cases; when this threshold is reached, *pfhrp2* deleted parasites are seeded into neighbouring regions such that neighbouring regions reach 1% genotype frequency after one year. Once a region reaches 1% genotype frequency, the future trajectory of deletions in that region is solely determined by the selection coefficient estimated for the region for a given parameter set. Given the use of a single fixed selection coefficient for each region, this assumes that malaria prevalence and case management in each region remains constant over time. Using this approach, we simulate a range of possible timelines for *pfhrp2* deletions in Africa.

## Results

### Parameter Estimation

In our analysis of the WHO Malaria Threat Maps database of *pfhrp2/3* deletions, we estimate that globally 63.5% (95% Confidence Interval: 55.1% - 71.1%) of *pfhrp2* deleted samples also had *pfhrp3* gene deletions. The distribution across studies of the percentage of *pfhrp2-*deleted samples with *pfhrp3* gene deletions was highly overdispersed, with significant differences between countries. Focusing on studies conducted in Africa (**Figure 2**), we estimate that 61.2% (95% CI: 47.9% - 72.7%) of *pfhrp2*-deleted samples also had *pfhrp3* gene deletions (**Figure 2A**). Further strong indication of the positive linkage between *pfhrp2* and *pfhrp3* deletions was observed based on the normalised coefficient of linkage disequilibrium, equal to 0.762 (see **Table 2**).

**Figure 2.**
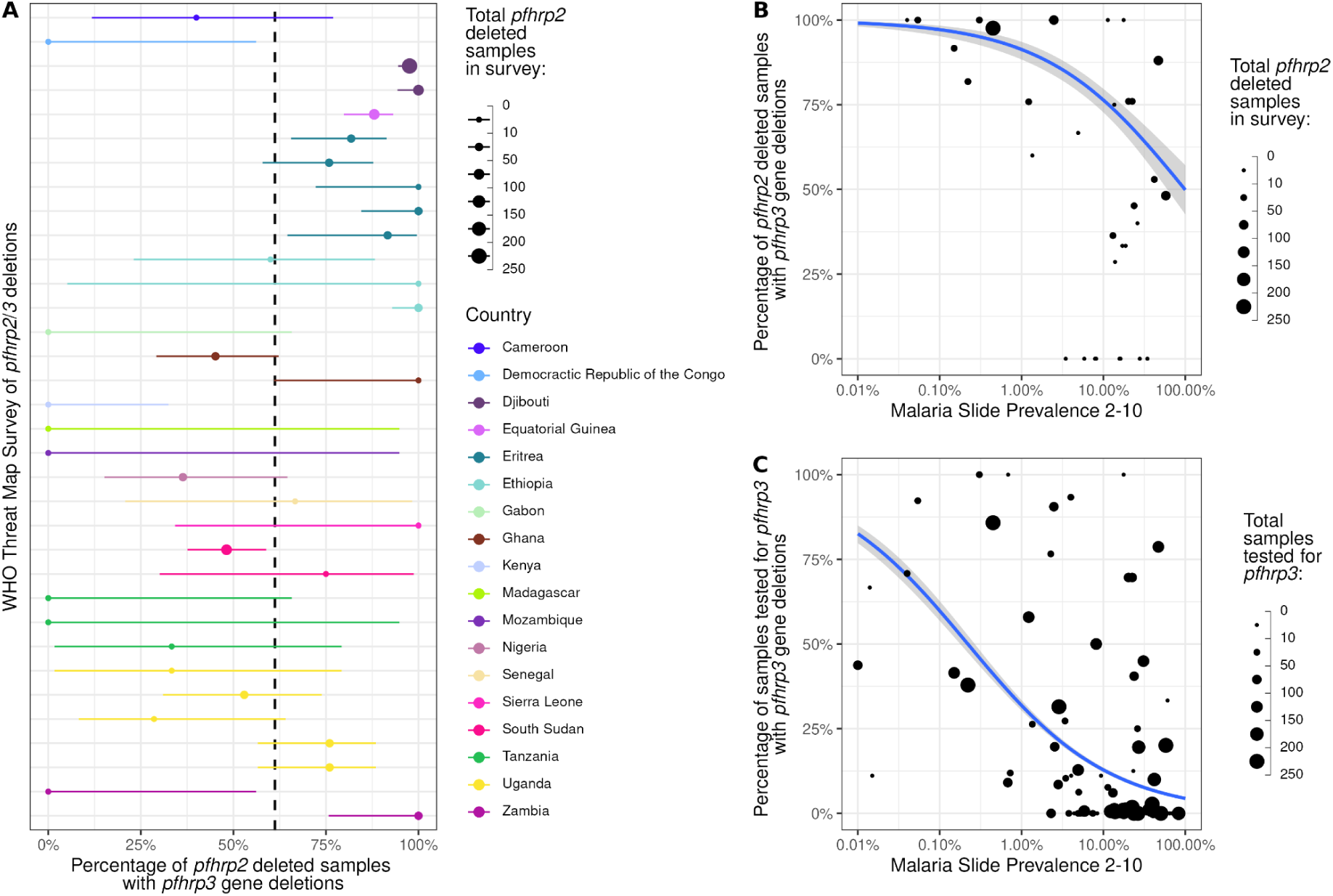
Distribution and independence of *pfhrp2/3* deletions in Africa collated in the WHO Malaria Threat Maps database. A) Percentage of *pfhrp2-*deleted samples also with *pfhrp3* deletions by survey. The mean and 95% confidence interval is shown with points and ranges. B). Relationship between the percentage of *pfhrp2-*deleted samples with *pfhrp3* deletions and malaria slide prevalence in 2-10 year-olds based on Malaria Atlas Project estimates. Binomial regression model fit (blue) shows the mean relationship between malaria prevalence and *pfhrp2* deletion frequency among *pfhrp3*-deleted parasites, with the 95% confidence interval of the regression fit shown with shaded bands. C). Relationship between the percentage of samples with *pfhrp3* deletions and malaria prevalence. In all plots, the point size represents the number of samples from each survey used to derive estimates.

**Table 2.**
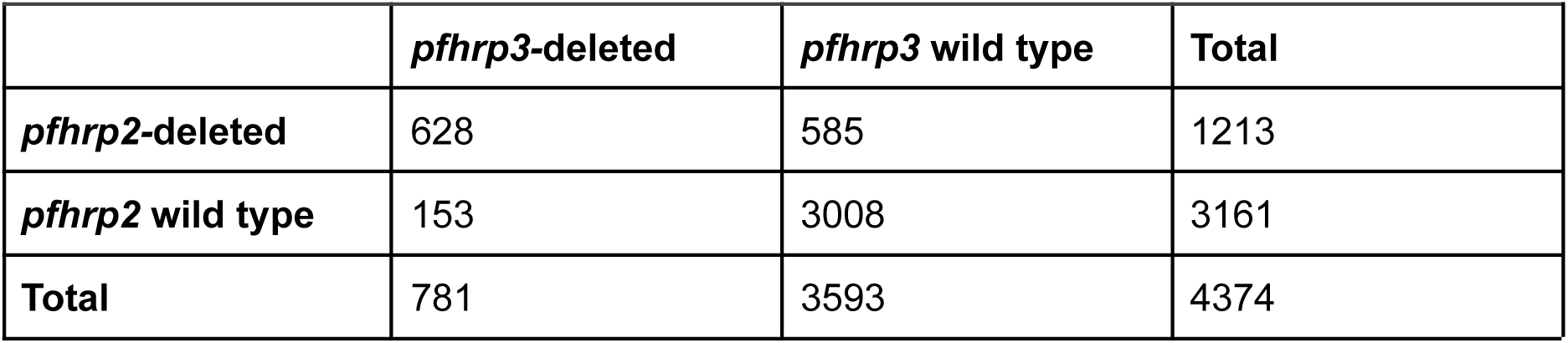
Frequency of *pfhrp2/3* deletions in Africa.

We find a significant relationship with malaria prevalence (X-squared = 1454.5, df = 1, p-value < 2.2e-16) (**Supplementary Table 1**), with surveys conducted in regions with higher malaria prevalence less likely to observe *pfhrp2-*deleted samples among samples with *pfhrp3* deletions (**Figure 2B**). We also observed significantly lower frequencies of *pfhrp3* deletions in surveys conducted in regions with higher malaria prevalence (**Figure 1B**). A different relationship between *pfhrp2/3* independence and malaria prevalence was observed on the other continents; studies in Asia showed insignificant associations between *pfhrp3* deletion frequency and malaria prevalence (**Supplementary Figure 1**).

In our model fitting exercise to jointly infer parameter values for both fitness costs and the cross reactivity of HRP3 epitopes, we estimate relative fitness of 96.4% (95% CrI: 95.8% - 97.0%) for *pfhrp2* deletions (i.e. deleted parasites relative contribution to onward infections each day is equal to 96% of that from wild-type parasites). Analogously, we estimate the probability that an infection due to only *pfhrp2* deleted parasites would still produce a positive HRP2-based RDT (HRP3 cross reactivity producing a positive test outcome) of 29.0% (95% CI: 20.0% - 45.0%) (see **Supplementary Figure 2**).

### Mapping the risk factors and modelling the impact on selection for *pfhrp2/3* deletions

We found parameter estimates for the majority of the risk factors identified, however, sources from reported WHO national data or the academic literature failed to identify suitable estimates for all malaria-endemic countries. To address this, we identified previous efforts by other groups that produced modelled parameter estimates at either the national or first administrative unit, notably the Commodities Forecast Dashboard by the Malaria Atlas Project (*25*). When compared against our literature review, we found broad agreement in the data sources identified (Supplementary Appendix 2), which resulted in similar estimates as produced by the Malaria Atlas Project for modelling trends in malaria commodities (*36*). However, we identified a number of outliers, totalling less than <0.5% of all parameters collected. These included outliers that reflected gaps in nationally reported data, e.g. zero reported cases to the WHO of malaria tested by RDT, and edge cases, such as ∼100% of care-seeking malaria infections who are not tested receiving treatment. In response, outliers were identified and multiple imputation using random forests was used to correct outliers based on the other collected covariates, yielding global maps of each parameter (**Supplementary Figures 3-6**).

Across model simulations that captured the full range of parameters identified for each country, we identified malaria prevalence as the most important determinant of the selection of *pfhrp2* deletions (**Supplementary Figure 7**). This is based on analysis of partial dependence of the statistical models used to predict selection coefficients, each of which provided unbiased predictions of selection coefficients (**Supplementary Figure 8A**) and exhibited strong predictive accuracy with less than 0.1 mean absolute error (**Supplementary Figure 8B**). We estimate that malaria prevalence has the greatest impact on selection coefficients when marginalising across all other parameters (**Supplementary Figure 7**), notably increasing at malaria prevalence less than 20% based on microscopy slide prevalence in 2-10 year olds (**Supplementary Figure 7A**). Treatment cascade parameters (non-adherence to RDT test outcomes, use of non-HRP2-based RDTs for testing and the HRP3 cross-reactivity) had similar inferred effect sizes (**Supplementary Figure 7 C-F**), reflecting their similar effects in altering the probability that an individual is only treated based on the outcome of an HRP2-based RDT.

### Innate and Prospective Risk of *pfhrp2* deletions

In scenarios in which each region has 1% of clinically relevant infections misdiagnosed due to *pfhrp2/3* gene deletions, we estimate that 73/106 countries modelled have at least one first administrative unit predicted to reach the 5% threshold within 20 years (**Figure 3**). We predict that the majority of the highest risk regions are very low transmission regions (<0.05% malaria prevalence), however, evolutionary trajectories in these settings are highly uncertain. The very low malaria prevalence and consequently small effective population size is predicted to increase the stochasticity in the dynamics of *pfhrp2* deletions - similarly to classical findings of the relationship between genetic drift and selection (*37*). Consequently, there is also the increased chance that deleted strains will stochastically fade-out due to small malaria population size, rather than increasing despite conditions being favourable for the selective advantage conferred by *pfhrp2* deletions to be realised. Conversely, we predict low risk of *pfhrp2* deletions in the highest malaria prevalence regions in Central and Western Africa, with estimated times to reach the 5% threshold in excess of 40 years.

**Figure 3.**
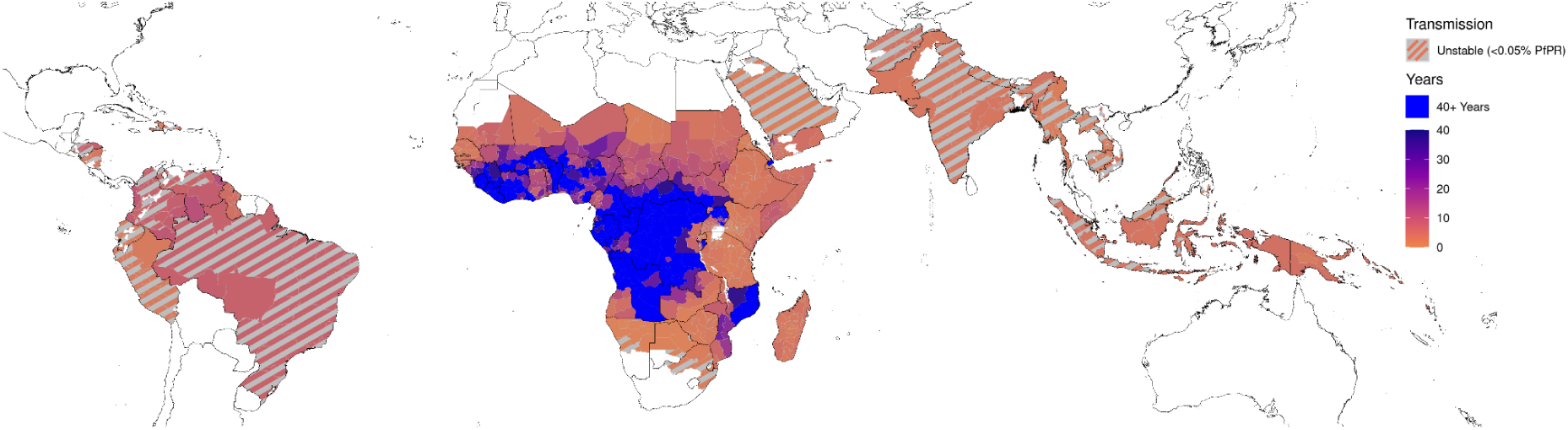
Global distribution of predicted times for the percentage of clinically relevant infections misdiagnosed due to *pfhrp2/3* gene deletions to increase from 1% to 5%. Regions estimated not to reach 5% within 40 years are shown in blue. Regions with very low, unstable malaria transmission (defined as <0.05% malaria prevalence) are shown with diagonal grey lines. (See **Supplementary Figure 9** for focus on Africa).

Focusing on countries with greater than 0.05% estimated malaria slide prevalence in 2020, we identify 20 countries in which the majority of first administrative units are classified as High Innate Risk (reaching the 5% threshold within 6 years) (see **Table 2**). All but three countries (Solomon Islands, Papua New Guinea and Guyana) are in Africa, with the majority of these countries in Africa representing those in which *pfhrp2/3* deletions have already been identified (e.g. Djibouti, Eritrea, Ethiopia, Gambia). However, we find a large range in assigned risk scores when we compare risk scores across the range of parameter uncertainty for each region (**Figure 4**). The majority of the uncertainty in selection speed for *pfhrp2* deletions is due to wide uncertainties in malaria prevalence for each administrative region. For example, malaria prevalence estimates in Yobe, Nigeria for 2020 range between 10% - 40%, which corresponds to an absolute change in selection coefficient of 0.3 (i.e an absolute increase of 30% in the annual proportional change in *pfhrp2* deletions). This change in predicted selection coefficients would result in a change in regional classification from Marginal concern (1% to 5% in >20 years) to High concern (1% to 5% in <6 years). Despite this uncertainty, we predict a number of regions that are consistently classified as High Concern, such as in Eritrea, Ethiopia, Zambia and Tanzania, and a number of regions in Central and West Africa that are consistently classified as Marginal risk (1% to 5% in >20 years).

**Figure 4.**
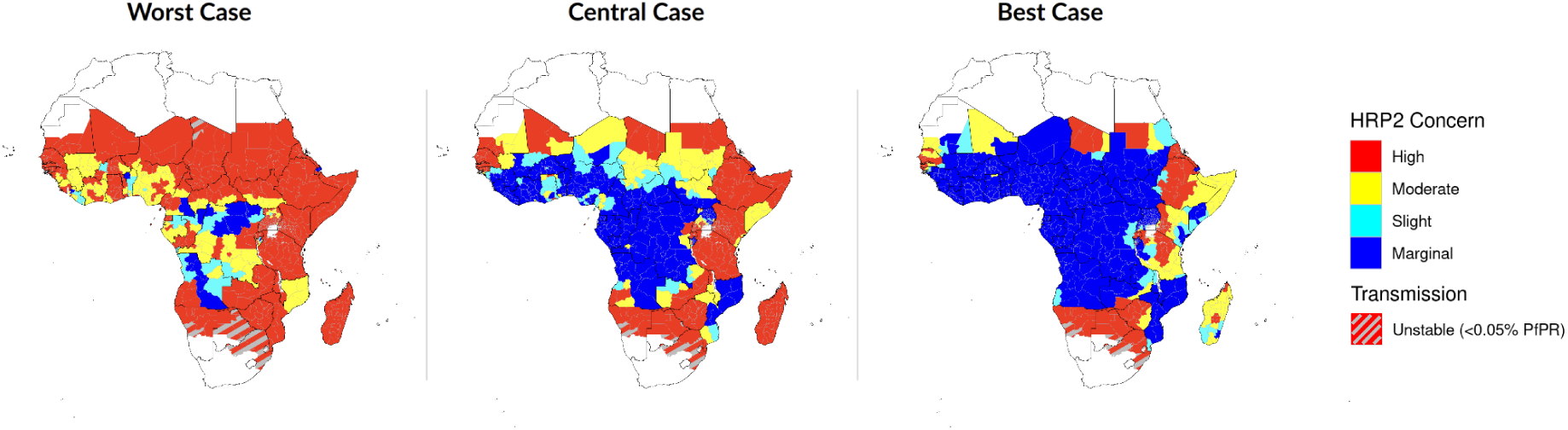
Innate risk score for the concern caused by *pfhrp2* deletions in Africa. High (red), moderate (yellow) and slight (teal) risk represent >5% of clinically relevant infections misdiagnosed due to *pfhrp2/3* gene deletions in less than 6, 12 and 20 years respectively, and marginal risk (blue) represents <5% in 20 years. Uncertainty in model parameters for each region impacts risk scores, with the worst and best case scenarios (based on the uncertainty in the range of parameters explored) shown. Regions with very low, unstable malaria transmission (defined as <0.05% malaria prevalence) are shown with diagonal grey lines. (See **Supplementary Figure 10** for global risk scores).

In scenarios in which we model the continued spread of *pfhrp2* deletions in Africa based on current estimates from the WHO Malaria Threat Maps, we predict that 29/49 of countries modelled have at least one first administrative unit predicted to reach the 5% threshold or have already reached the 5% threshold within 20 years (**Figure 5**). If HRP2-based RDTs remain the mainstay of malaria case management, we predict that the major route for *pfhrp2* deletions to spread south out from the current hotspot in the Horn of Africa, moving through East Africa over the next 20 years. Additionally, deletions identified in Western Africa are predicted to increase, especially in Senegal and Mali. Prospective risk scores classify fewer regions as high risk than Innate risk scores (**Supplementary Figure 11**). Across both risk scores, however, a number of countries are predicted to be majoritively (more than 50% of first administrative units) identified as being High Risk (**Table 3**), including Djibouti, Eritrea, Ethiopia, Senegal, Zambia and Kenya. Similar to the Innate risk score, there is considerable uncertainty in the modelled timelines for the spread of deletions. Interactive risk maps for each parameter scenario are available at https://worldhealthorg.shinyapps.io/DeletionRiskExplorer (see **Supplementary Figure 12).**

**Figure 5.**
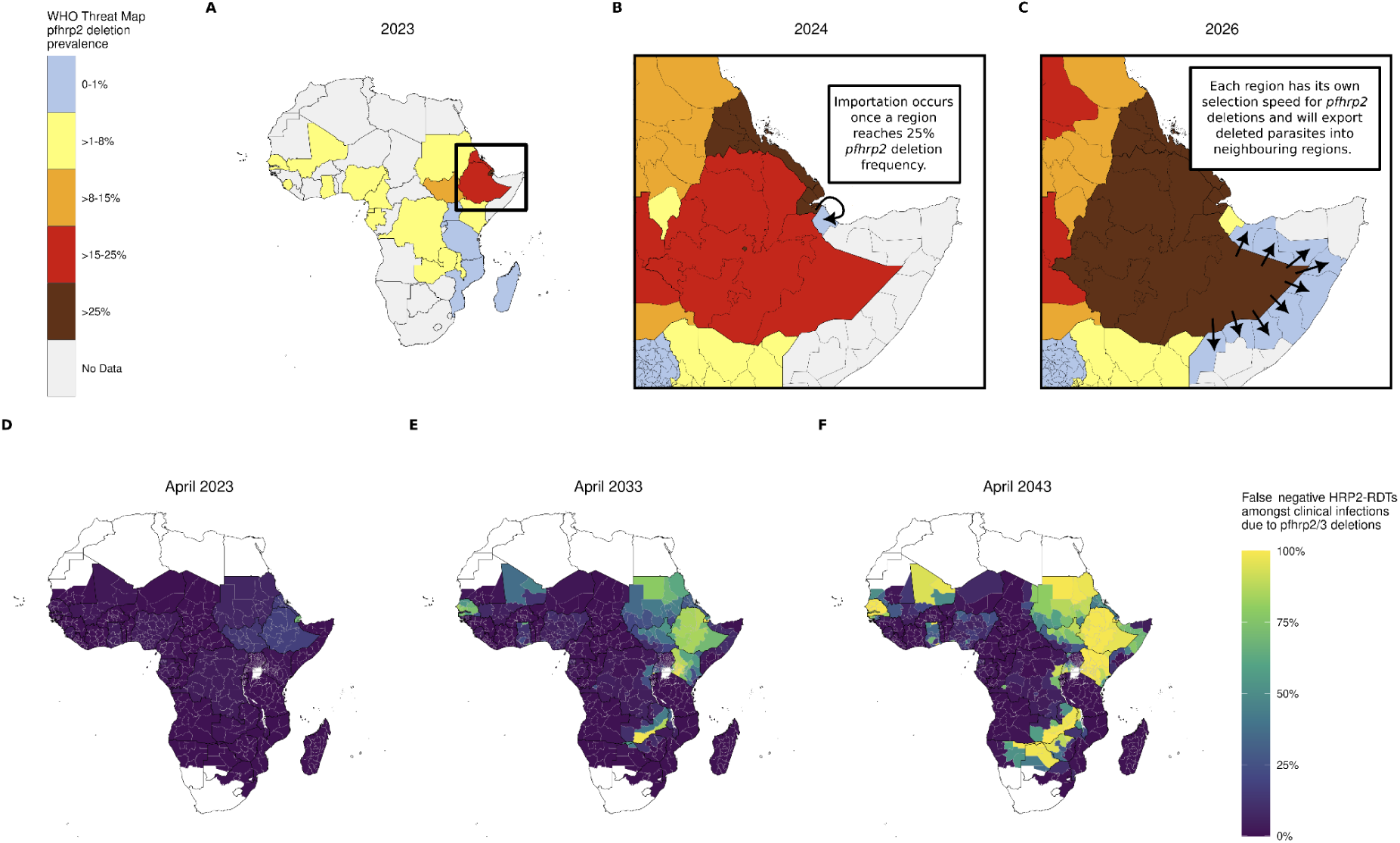
Prospective risk scores for *pfhrp2* deletions in Africa. The Prospective risk score models continued spread of deletions based on current best estimates of the prevalence of *pfhrp2* deletions as collated in the WHO Malaria Threat Maps database (shown in A). In this model, we make the assumptions that B) deletions are imported into a region from a neighbouring region once they reach a prevalence of 25%, and that C) selection of deletions in a region is determined by that region’s transmission intensity and treatment-related parameters.D) - F) show the predicted spread of false-negative RDTs due to *pfhrp2/3* deletions in Africa over the next 20 years (see Supplementary Video 1).

**Table 3.** High risk countries by risk score. Percentage of first administrative regions classified as high **A)** Innate or **B)** Prospective risk (>5% of clinically relevant infections misdiagnosed due to *pfhrp2/3* gene deletions in less than 6 years given a starting frequency of *pfhrp2* deletions of 1%) is shown. Only countries in which 50% or more regions are classified as high risk are shown.

| Country | % Admin 1 regions with high Innate risk |
| --- | --- |
| Eritrea | 100.0% |
| Ethiopia | 100.0% |
| Gambia | 100.0% |
| Madagascar | 100.0% |
| Namibia | 100.0% |
| Papua New Guinea | 100.0% |
| Rwanda | 100.0% |
| Senegal | 100.0% |
| Tanzania | 100.0% |
| Zimbabwe | 100.0% |
| Solomon Islands | 100.0% |
| Comoros | 100.0% |
| Kenya | 93.6% |
| Guinea-Bissau | 88.9% |
| Yemen | 84.2% |
| Guyana | 80.0% |
| Mauritania | 66.7% |
| Djibouti | 60.0% |
| Somalia | 50.0% |
| Zambia | 50.0% |

**B)**
| Country | % Admin 1 regions with high Prospective risk |
| --- | --- |
| Djibouti | 100.0% |
| Eritrea | 100.0% |
| Ethiopia | 100.0% |
| Senegal | 100.0% |
| South Sudan | 100.0% |
| Sudan | 100.0% |
| Kenya | 95.7% |
| Ghana | 90.0% |
| Equatorial Guinea | 85.7% |
| Zambia | 60.0% |

## Discussion

In this study, we model the global risk of selection and spread of *pfhrp2* deletions and confirm a significant threat to malaria control efforts in Africa if case management continues to rely upon HRP2-based diagnosis. Incorporating the most recent understanding of deletions and the best estimates of key model parameters, we find that malaria prevalence is the most important driver of deletions globally. However, uncertainty in malaria prevalence data, further exacerbated by the pandemic-induced delay in key data sources such as Demographic and Health Surveys, limits confidence in regional risk estimates. In response, we investigated a range of scenarios and uncertainties to identify countries and regions at highest risk from deletions across the range of scenarios explored. Globally, most malaria-endemic areas and especially those with very low prevalence are predicted to select for deletions rapidly. In Africa, this includes regions in the Horn of Africa, East Africa, and a few countries in West Africa, such as Senegal and Mali.

Our findings contrast with earlier *pfhrp2* deletion risk maps and timelines (*17*) in several significant ways. First, our approach focuses on a different outcome measure, namely the proportion of clinically relevant malaria cases misdiagnosed due to gene deletions, consistent with current WHO policy guidance (*30*). Second, we incorporate the best available data on current deletion prevalence to evaluate how deletions may spread between regions Third, we produce an interactive tool for decision makers to explore the risk maps for each parameter scenario and understand how each parameter impacts the selection of *pfhrp2* deletions. However, despite incorporating current best estimates, these projections need to be viewed with the appropriate uncertainty due to considerable gaps in surveillance of *pfhrp2/3* deletons as well as heterogeneity in the quality and consistency of previously conducted *pfhrp2/3* surveys (*7*). Consequently, the results should be viewed as tools to consider how the two components for mapping the potential spread of deletions - a region’s innate susceptibility for deletions to increase once established (dependent on a region’s malaria transmission intensity, treatment-seeking data, and RDT usage data) and the spatial connectivity to regions with high levels of deletions - may interact to drive the spread of deletions. Despite their simplicity, these results can help guide control interventions to stem the threat of *pfhrp2/3* deletions, particularly in identifying regions that need to be prioritised for surveillance to provide accurate data before deciding whether to switch front-line RDTs. Outside of regions that have already switched front-line RDTs, these include Senegal, Zambia and Kenya.

Fewer regions are identified as High risk based on the Prospective risk score compared to the Innate risk score for two primary reasons. First, the Prospective risk score incorporates estimates of the proportion of RDTs in use in a country that are not only HRP2-based. Consequently, countries that primarily use non-HRP2-based RDTs, such as Rwanda (primarily using Pf/PAN RDTs based on Global Fund and President’s Malaria Initiative data), will not select for *pfhrp2* deletions. Second, the Prospective risk score is seeded with current estimates of *pfhrp2* deletion prevalence in each country. Countries without surveys or less than 1% *pfhrp2* deletions, such as Tanzania, are predicted to reach the 5% threshold slower than in the Innate risk scenario, which explores timelines from a starting frequency of 1% *pfhrp2* deletions. We chose to produce two risk maps (the Innate and Prospective risk) because robust molecular surveys of *pfhrp2/3* deletions have not been conducted across all regions. Although surveillance for *pfhrp2/3* deletions has increased rapidly since the widespread introduction of RDTs, by the beginning of 2023, surveys have only been conducted in 22 countries in Africa (*21*). For the Prospective risk score, we made the simplifying assumption that countries without surveys have 0% *pfhrp2* deletion frequency. If this assumption is incorrect, the Prospective score will underestimate the risk in these countries. The Innate risk score, on the other hand, simply focuses on the risk that *pfhrp2* deletions pose once present in a region (and assuming the region has not switched to non-HRP2-based RDTs alone or in combination with HRP2). This dual approach has several advantages. The Innate risk score can be used to confirm that the model correctly identifies regions in which deletions have rapidly increased as High risk. Indeed, the maps of Innate risk (**Figure 3**) correctly identify the Horn of Africa as a region of consistent high risk. The Innate risk score can also be used to address additional questions relevant to malaria policies, including where to prioritize surveillance given plateauing levels of funding and competing demands (*1*). For example, if deciding amongst countries without previous surveys, the Innate risk score can be used to identify countries predicted to select for deletions fastest and therefore in greatest need of surveillance and/or early transition to non-HRP2-based RDTs.

Our approach has several important limitations. First, our exploration of international spread employs a simplistic approach for how deletions are exported between regions. Second, the model parameters carry a high degree of uncertainty. Our estimates of fitness costs are derived from model fitting to a handful of surveys, and may not be reflective of the fitness costs associated with *pfhrp2* deletion in parasites outside of the Horn of Africa. Once additional longitudinal deletion data is available, selection coefficients can be more accurately inferred, and fitness costs can be better estimated. However, the degree of uncertainty in certain key parameters, such as malaria prevalence, highlights the need for data to provide more precise estimates of key drivers of *pfhrp2/3* selection. These same data are needed to model the spread of artemisinin partial resistance (*31*), which is now spreading in a number of regions in Africa (*38–40*). Third, our model assumes that malaria prevalence and treatment remain constant in the future. Fourth, the country-specific estimates of linkage between *pfhrp2* and *pfhrp3* deletions provided here assume that the dynamics of these two loci are at equilibrium and that no selective forces are acting to pull certain genotypes, such as *pfhrp2-/pfhrp3-*, to higher levels. However, we have observed a significant relationship between deletions and malaria prevalence that aligns with recent mechanistic explanations of how *pfhrp3* deletions arise and may be driven by low malaria prevalence (*22*). If malaria prevalence falls in a region, in addition to the increased selection of *pfhrp2* deletions that occurs at low prevalence, the frequency of *pfhrp3* deletions may also increase furthering the selection of *pfhrp2* deletions. In response, additional surveillance data of both *pfhrp2* and *pfhrp3* deletions is needed, which can be leveraged to test hypotheses of how non-RDT mediated processes drive *pfhrp3* deletion emergence and subsequently create a selective niche for *pfhrp2* deletions. Lastly, while we have modelled how HRP2-based RDTs create a selective pressure for *pfhrp2* deletions, this process does not capture the historic process by which *pfhrp2* deletions have emerged in South America, which occurred without this pressure. These results are, however, still relevant in identifying that these regions are susceptible to selecting for deletions given the low malaria prevalence if they relied on HRP2-based RDTs, while also noting that a greater understanding of the fundamental biology and evolution that led to the selection of *pfhrp2* deletions in regions in South America is needed.

The issues surrounding spread of *pfhrp2/3* deletions are not unique to malaria. Management strategies for controlling RDT-evasive genotypes can be borrowed from the drug-resistance management literature which provides evaluations of how multiple antimalarial therapies can be deployed (*41*, *42*). RDTs employing multiple proteins for diagnosis (e.g. PfHRP2 and PfLDH) are analogous to combination therapies in that a parasite lineage would need to acquire two genetic mechanisms simultaneously to evade detection. Deployment of both HRP2-based RDTs and non HRP2-based RDTs in a single population is similar to the multiple first-line therapies (MFT) (*43*) approach of slowing down resistance in that an RDT-evasive parasite is likely to undergo diagnosis with a different RDT in the next patient it infects. These approaches would first need to be field tested to ensure adequate procurement, distribution, and compliance before evaluating their potential for slowing down or reversing the evolution of RDT evasion.

In conclusion, this study provides a refined and updated prediction model for the emergence of *pfhrp2/3* deletions. Despite its limitations, our models offer valuable insights that can help policymakers prioritize surveillance and future deployment of alternative RDTs, leveraging our interactive tool to identify the regions that are consistently identified as high risk. It also should signal to test developers and manufacturers where new markets are likely to emerge first for alternatives to exclusive HRP-RDTs. As our understanding of the complex processes driving *pfhrp2/3* deletions improves and more data become available, we will continue to refine and update our predictions and monitor the increasingly concerning threat posed by *pfhrp2/3* deletions.

## Data Availability

All source code, analysis, tables and plots are available at https://github.com/OJWatson/hrpupv0.2.0.

https://github.com/OJWatson/hrpup/releases/tag/v0.2.0

## Acknowledgements

RJZ, TN-AT, MFB are supported by the National Institutes of Health (NIAID R01AI153355) and the Bill and Melinda Gates Foundation (INV-005517). OJW is supported by an Imperial College Research Fellowship sponsored by Schmidt Futures. TS, SR, PAD and PG are supported by the Bill and Melinda Gates Foundation (INV-009390/OPP1197730).

