## Supplementary Materials for "Global risk of selection and spread of *Plasmodium falciparum* histidine-rich protein 2 and 3 gene deletions"

### Supplementary Material

*For the purpose of open access, the author has applied a 'Creative Commons Attribution (CC BY) licence (where permitted by UKRI, 'Open Government Licence' or 'Creative Commons Attribution No-derivatives (CC-BY-ND) licence' may be stated instead) to any Author Accepted Manuscript version arising.*

### Supplementary Figures

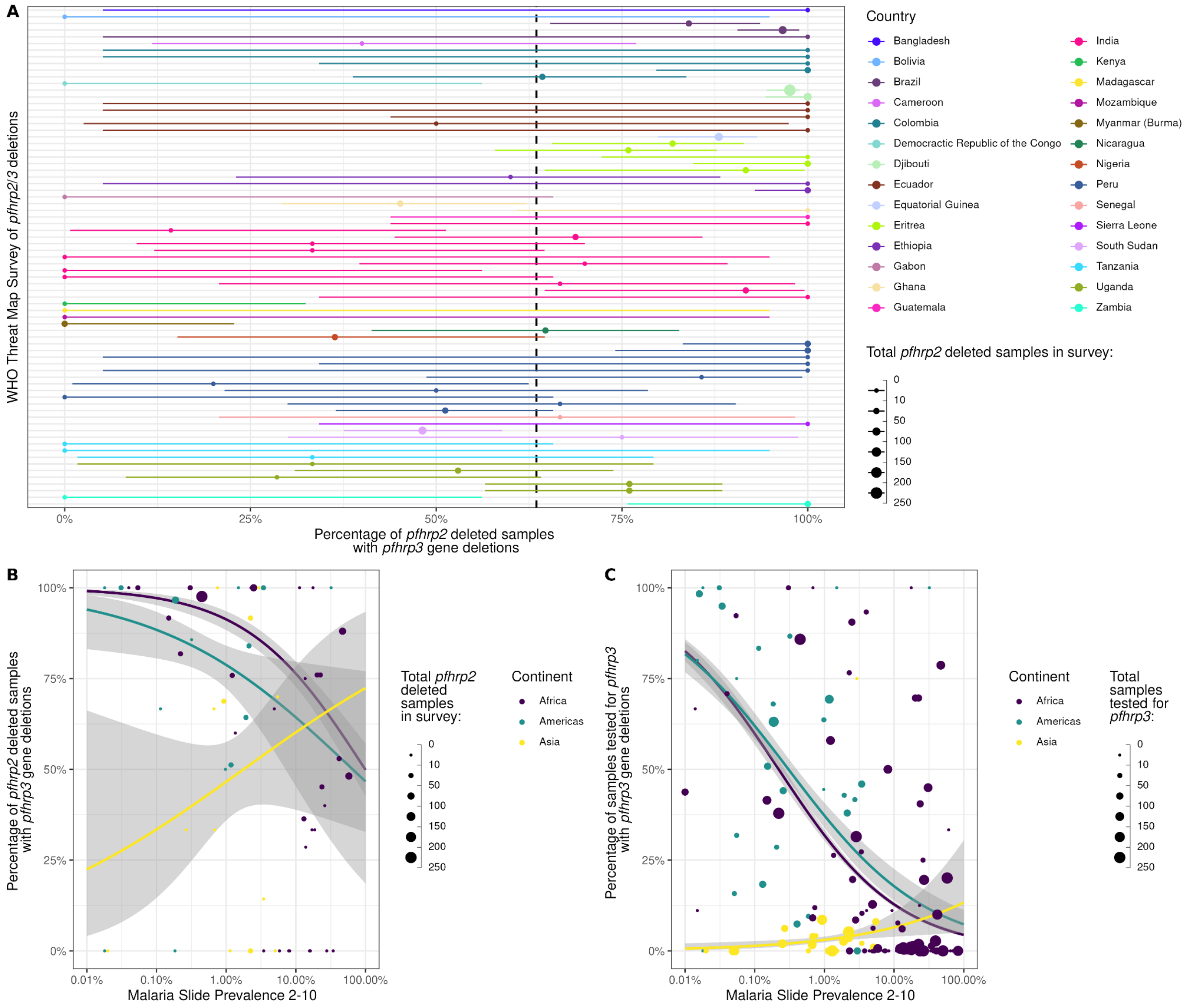


**Supplementary Figure 1.** Distribution and independence of *pfhrp2/3* deletions globally collated in the WHO Threat Maps database. A) Percentage of *pfhrp2*-deleted samples also with *pfhrp3* deletions by survey. The mean and 95% confidence interval is shown with points and ranges. B). Relationship between the percentage of *pfhrp2*-deleted samples also with *pfhrp3* deletions against malaria slide prevalence in 2-10 year-olds based on Malaria Atlas Project estimates. Binomial regression model fit (blue) shows the mean relationship between malaria prevalence and *pfhrp2* deletion frequency among *pfhrp3*-deleted parasites, with the 95% confidence interval of the regression fit shown with shaded bands. C). Relationship between the percentage of samples with *pfhrp3* deletions and malaria prevalence. In all plots, the point size represents the number of samples from each survey used to derive estimates. In both B) and C) the regression relationships are shown for each continent.

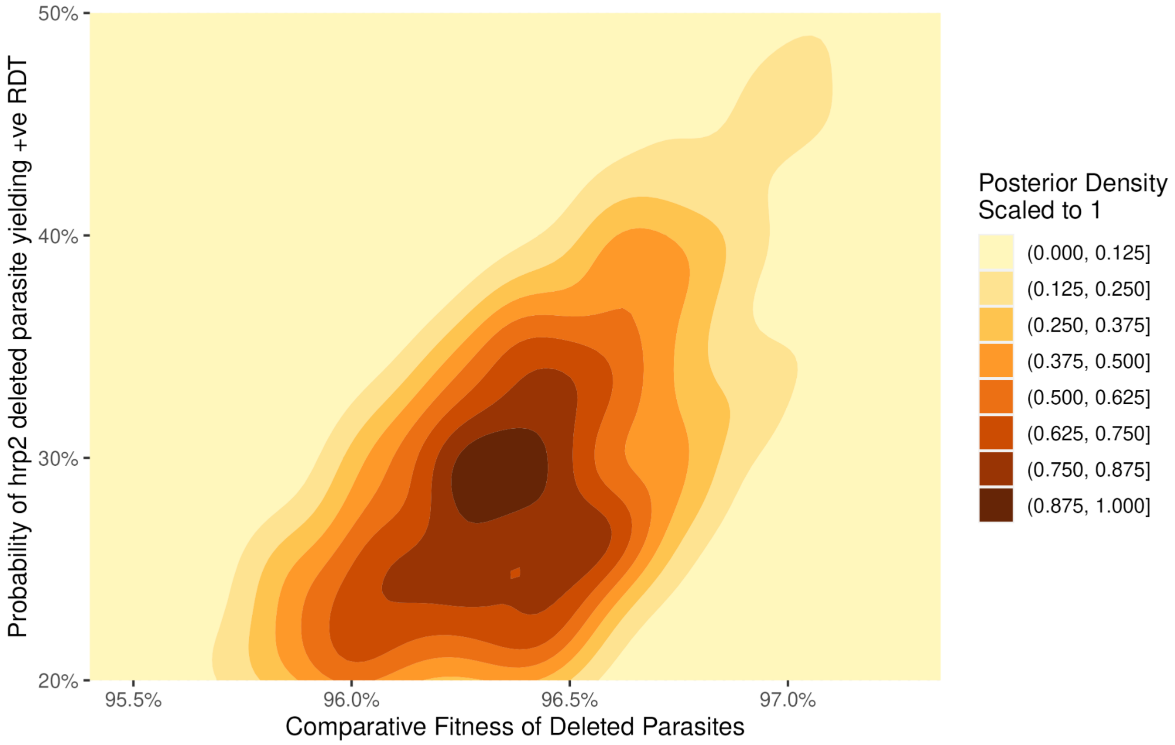


**Supplementary Figure 2.** Parameter estimation of fitness and HRP3 cross-reactivity. Selection trends for studies in Ethiopia and Eritrea reporting frequencies of *pfhrp2* deletions were created based on historical malaria prevalence and treatment seeking trends and spanning likely ranges for HRP3 cross reactivity and comparative fitness costs. 1000 parameter pairs were drawn from the posterior distribution and the resultant posterior density scaled to a maximum of 1 is shown, with darker colours showing a higher area of likelihood.

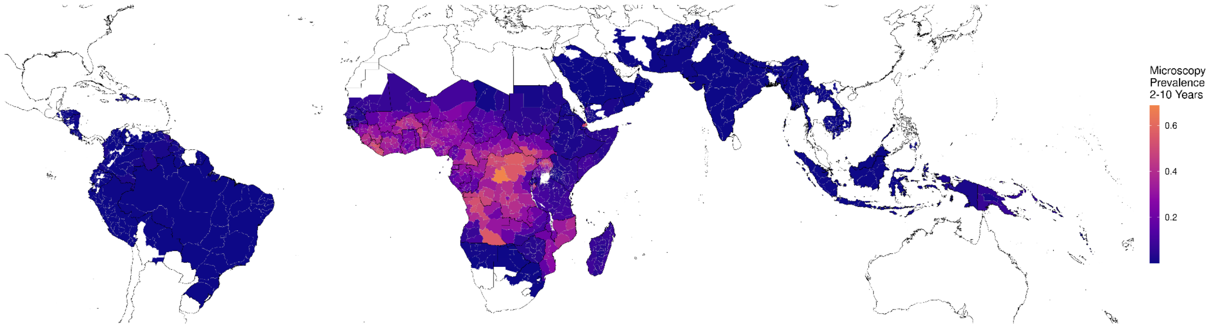


**Supplementary Figure 3.** Median malaria slide prevalence in children aged 2-10 in 2020 based on Malaria Atlas Project estimates and aggregated to the first administrative unit.

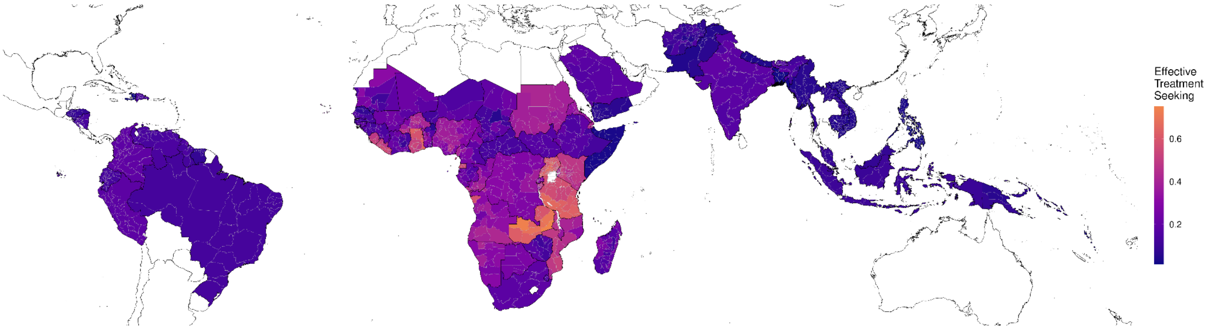


**Supplementary Figure 4.** Median effective treatment in 2020. Effective treatment seeking reflects the probability that an individual with symptomatic malaria seeks treatment and is subsequently treated. Estimates are shown at the first administrative unit.

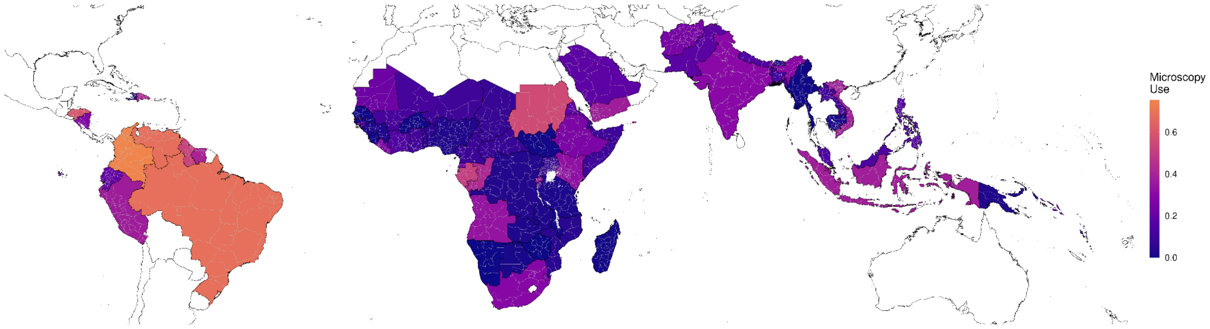


**Supplementary Figure 5.** Proportion of all diagnostic testing conducted using microscopy. Estimates available at the national level.

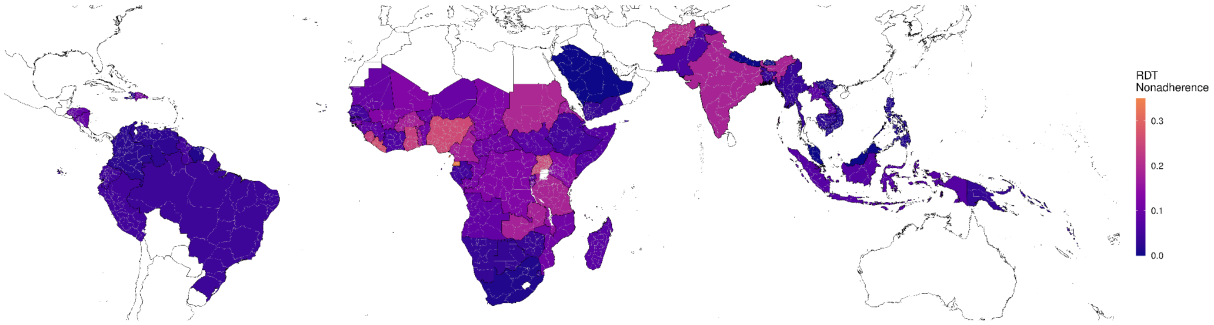


**Supplementary Figure 6.** Proportion of false-negative RDT tests that are subsequently treated for malaria, i.e. non-adherence to RDT test outcomes. Estimates available at the national level.

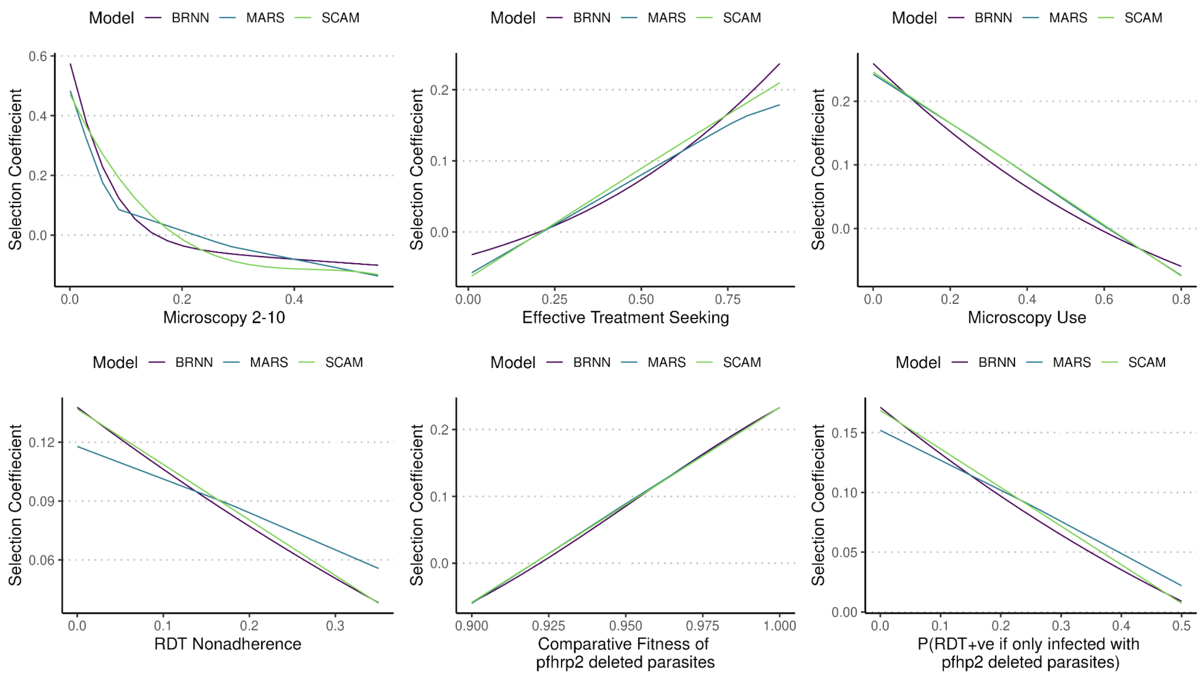


**Supplementary Figure 7.** Partial dependence plots of each individual model used in the ensemble model for predicting selection coefficients. Partial dependence is shown across the six model inputs that were explored when modelling the risk of *pfhrp2* deletions.

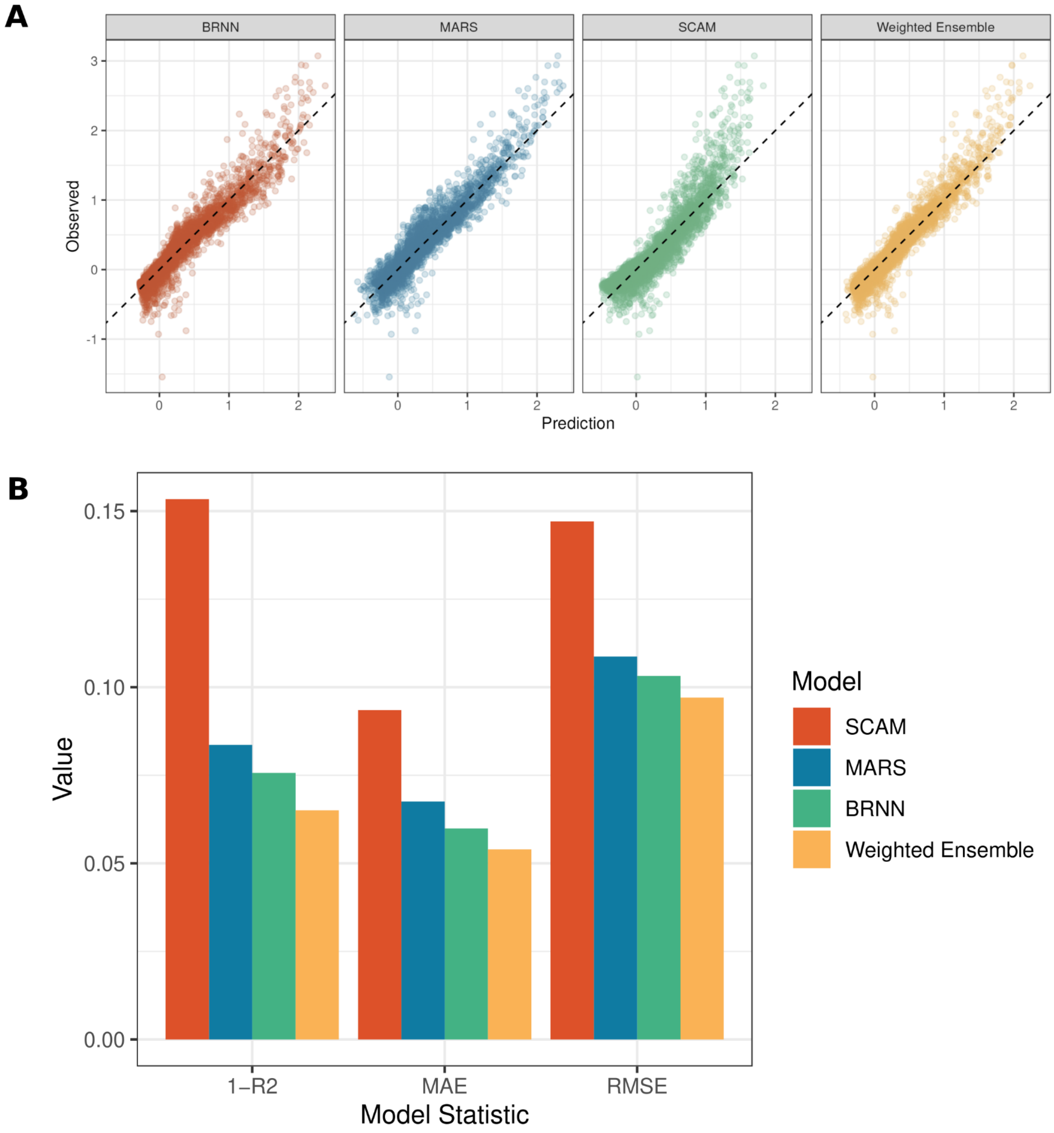


**Supplementary Figure 8.** Ensemble model fitting performance on hold-out test data. A) Model prediction selection coefficients are shown against the observed selection coefficients, with y = x trend line shown with a dashed line. B) Model performance summary statistics for each individual model alongside the weighted ensemble model.

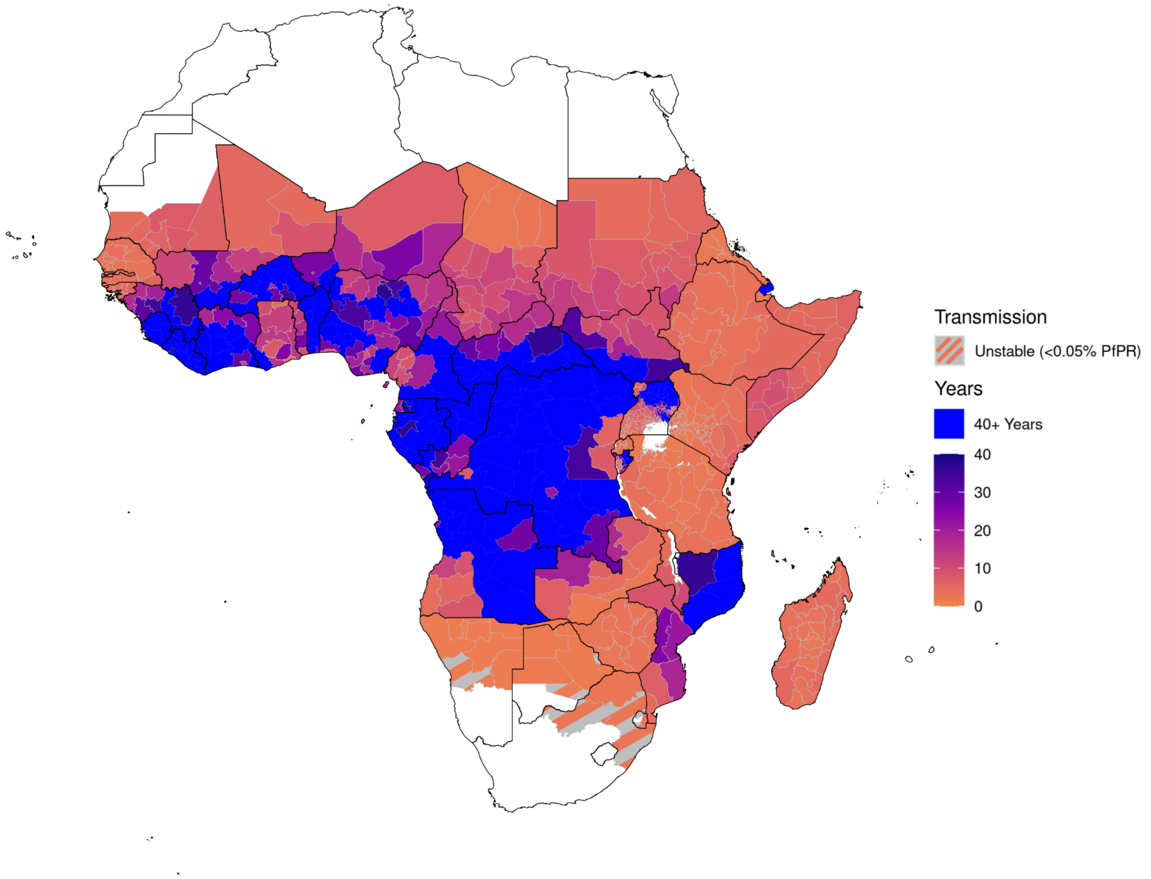


**Supplementary Figure 9.** Predicted central estimates for the time in years for the percentage of clinically relevant infections misdiagnosed due to *pfhrp2/3* gene deletions to increase from 1% to 5% in Africa. Regions estimated to not reach 5% within 40 years are shown in blue. Regions with very low, unstable malaria transmission (defined as <0.05% malaria prevalence) are shown with diagonal grey lines.

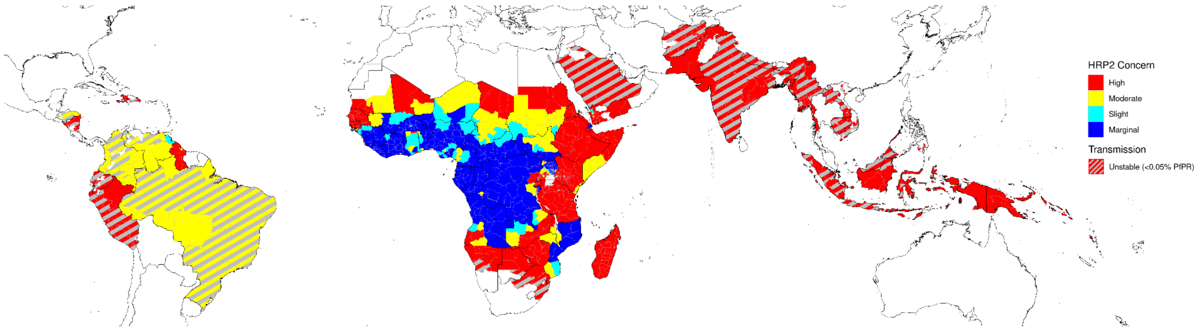


**Supplementary Figure 10.** Central estimates of the Innate risk score for the concern caused by *pfhrp2* deletions globally. High (red), moderate (yellow) and slight (teal) risk represent >5% of clinically relevant infections misdiagnosed due to *pfhrp2/3* gene deletions in less than 6, 12 and 20 years respectively, and marginal risk (blue) represents <5% in 20 years. Regions with very low, unstable malaria transmission (defined as <0.05% malaria prevalence) are shown with diagonal grey lines.

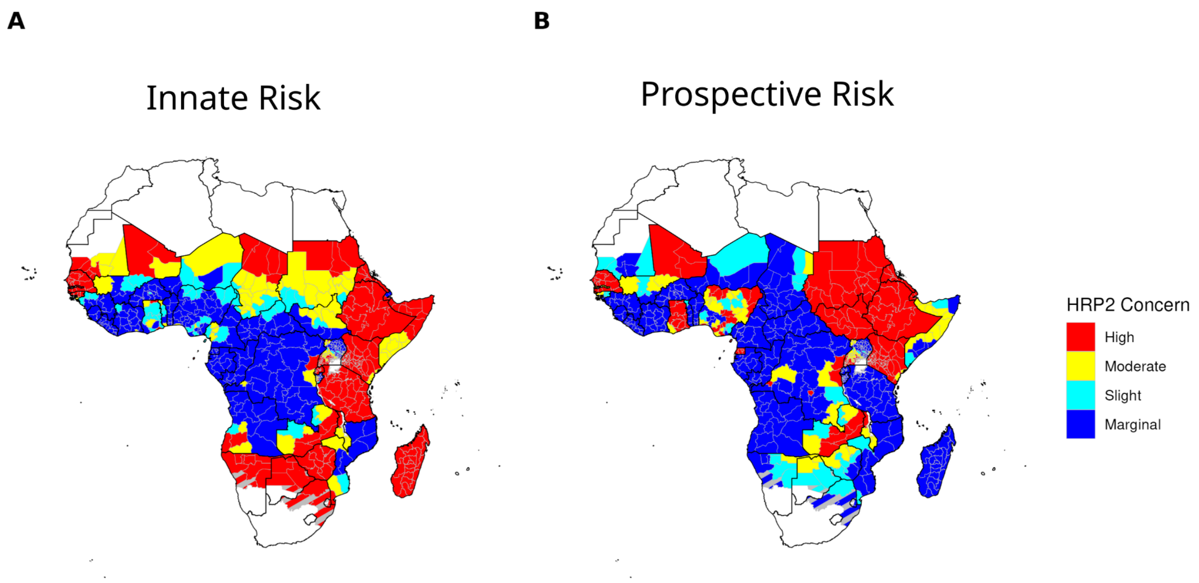


**Supplementary Figure 11.** Innate and Prospective risk scores for Africa. Risk scores are presented for our central estimates of covariates. High (red), moderate (yellow) and slight (teal) risk represent >5% of clinically relevant infections misdiagnosed due to *pfhrp2/3* gene deletions in less than 6, 12 and 20 years respectively, and marginal risk (blue) represents <5% in 20 years. Regions with very low, unstable malaria transmission (defined as <0.05% malaria prevalence) are shown with diagonal grey lines.

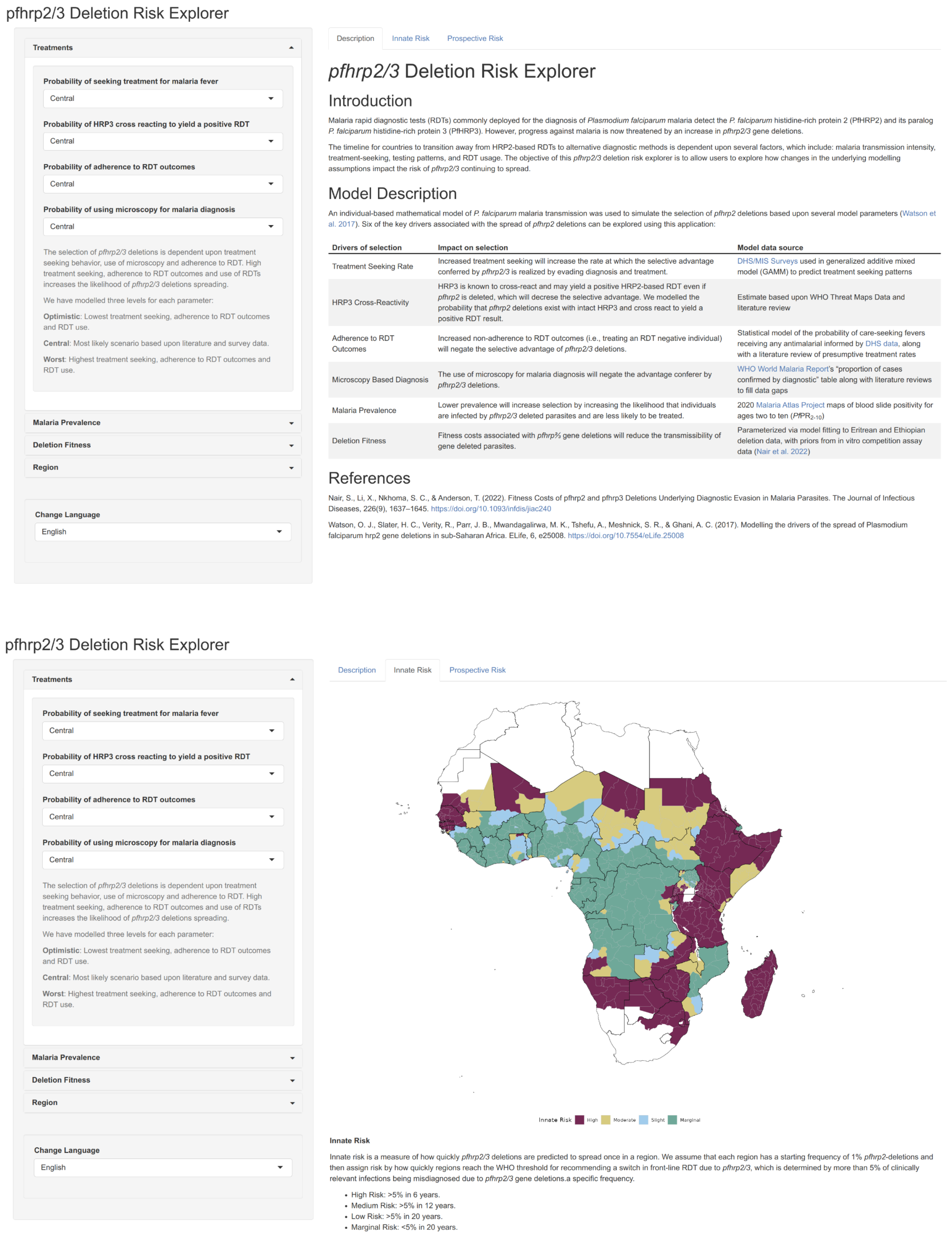


**Supplementary Figure 12.** Example interactive interface for exploring the full range of uncertainty in *pfhrp2* risk. The interface allows users to explore how changing scenarios explored (Worst, Central, Optimistic case scenarios with respect to *pfhrp2* deletions spread) impacts the risk posed by *pfhrp2* deletions.

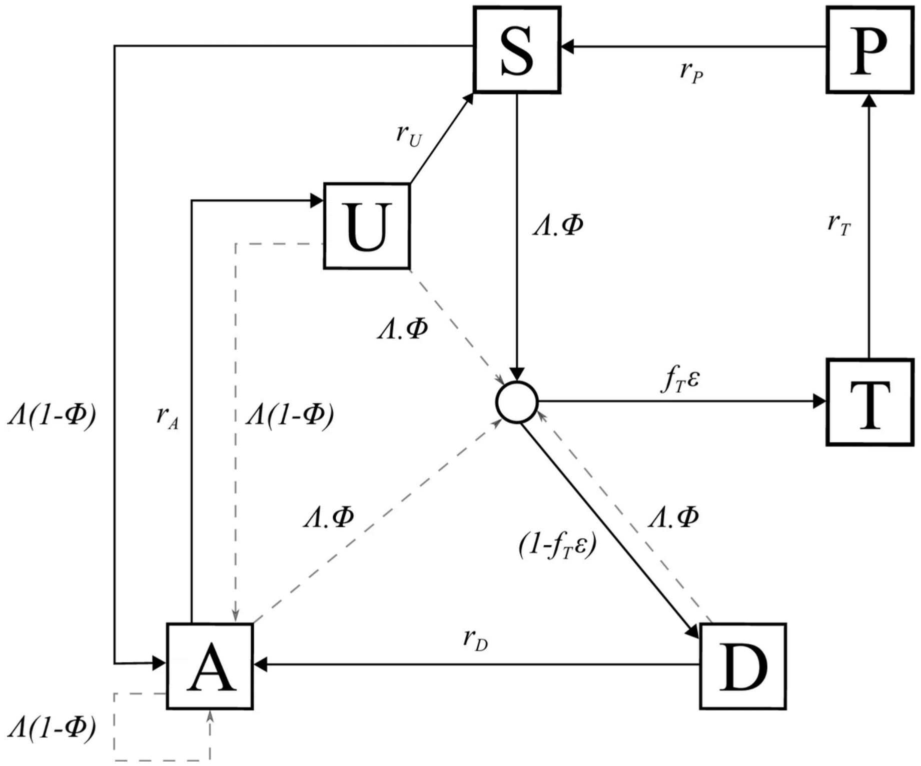


**Supplementary Figure 13.** Flow diagram for the human component of the transmission model, with dashed arrows indicating superinfection. S, susceptible; T, treated clinical disease; D, untreated clinical disease; P, prophylaxis; A, asymptomatic patent infection; U, asymptomatic sub-patent infection; $r_x$, rates of transition between compartments; $f_T$, effective treatment seeking (seeking treatment that will result in effective treatment unless diagnostic test results in a false-negative outcome due to *pfhrp2* deletions; $\Lambda$, force of infection; $\Phi$, probability of developing clinical symptoms that may trigger seeking treatment; $\varepsilon$, probability of HRP3 epitopes present and cross reacting to yield positive HRP2-based RDT.

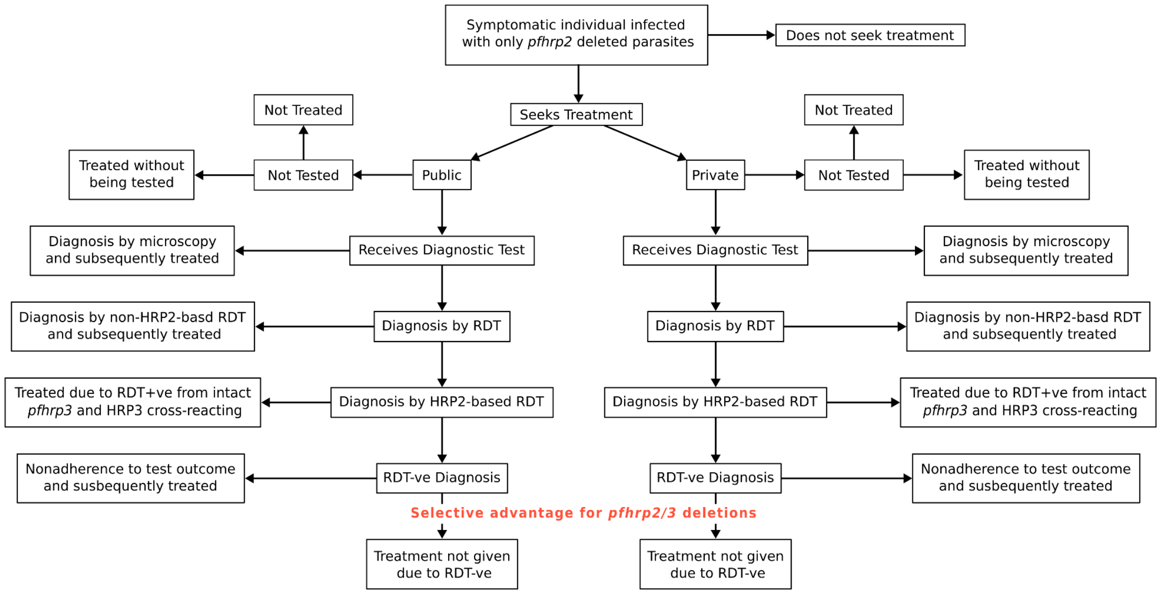


**Supplementary Figure 14.** Full treatment cascade pathways. Routes to treatment outcomes are aggregated when included in the transmission model, with parameters for each process determined from our parameter and literature review.

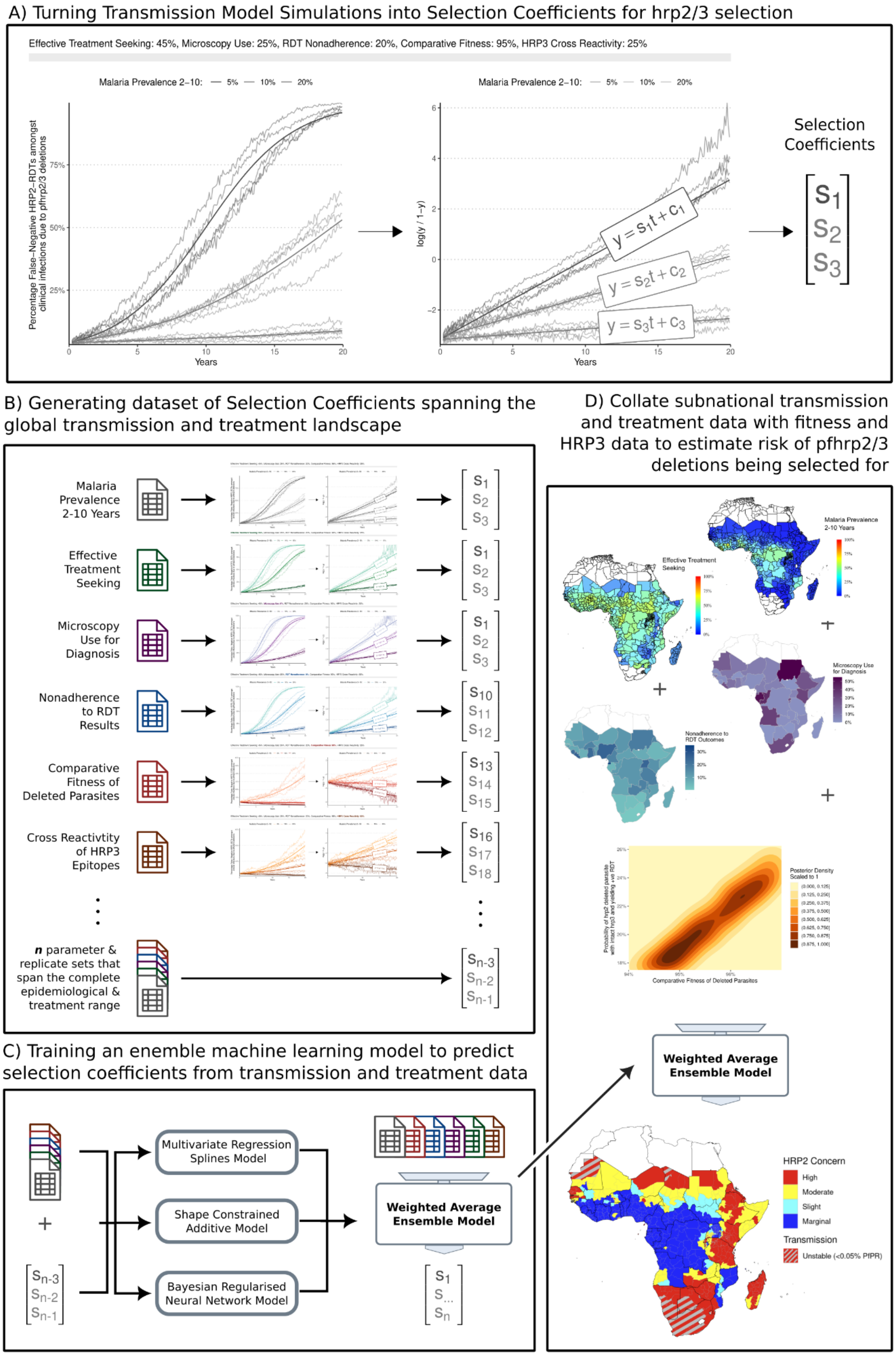


**Supplementary Figure 15.** Methodological schematic. A) Estimation of selection coefficients from stochastic model simulations. B) Generating training data using simulation studies that feed into C) a statistical model for estimating selection coefficients. D) The final model is used to estimate the risk posed by *pfhrp2/3* deletions using collated subnational covariates and parameters from model fitting exercises.

#### Supplementary Videos

Video 1. Time series animation of the spread of *pfhrp2* deletions in Africa based on prospective risk score modelling.

### Supplementary Tables

**Table 1.** Binomial modelling of the impact of malaria prevalence on *pfhrp2/3* status globally

| | *pfhrp3* gene deletion prevalence of *pfhrp2* deleted samples (95% CI) | *pfhrp3* gene deletion prevalence (95% CI) |
|---|---|---|
| Malaria Prevalence | -0.2892 (-0.3293, -0.2490) | -0.5502 (-0.5812, -0.5193) |
| | p: <2e-16 | p: <2e-16 |
| Observations | 177 | 193 |
| Log Likelihood | -2,044.82 | -3,014.87 |

### Supplementary Methods

#### Model parameters and Literature Review

Details of the literature review and comparisons between data sources is available in Supplementary Appendix 2. The review described the sources of data identified and results of a literature review to estimate parameters needed as inputs into the *pfhrp2* transmission model (Supplementary Figure 13), which are estimated by aggregating pathways from the typical treatment cascade (Supplementary Figure 14).

#### Ensemble Machine Learning Model for Predicting Selection Coefficients

We trained an ensemble machine learning model to predict selection coefficients estimated from model simulations that span the range of range of malaria prevalence and treatment coverages observed globally. Specifically, we conducted model simulations that varied six parameters: 1) the malaria prevalence; 2) the probability of an individual seeking treatment and being effectively treated after having received a diagnostic test; 3) the adherence to test outcomes for deciding on treatment; 4) the proportion of all diagnoses that occur using microscopy; 5) the relative fitness of *pfhrp2* deleted parasites; and 6) the probability that an individual only infected with *pfhrp2* deleted parasites yields a positive HRP2-based RDT due to the parasites not having a *pfhrp3* deletion and the resultant HRP3 cross-reacting with the RDT yielding a positive test.

We constructed 8748 unique sets of the 6 described parameters. For all parameter combinations, five stochastic realisations of 100,000 individuals were simulated for 40 years to reach equilibrium first before simulating the selection of *pfhrp2* deletions over the following 20 years, with a starting frequency of *pfhrp2* deletions equal to 6% (similar to our previous analysis (*1*). From each simulation we recorded the monthly proportion of clinically relevant infections that would be misdiagnosed due to *pfhrp2/3* gene deletions (i.e. clinical infections only infected with *pfhrp2* deletions and assumed to also have *pfhrp3* deletions). We subsequently calculated selection coefficients (the annual % change in proportion of misdiagnosed clinical cases) for each simulation repetition by linear regression of the log odds of a clinical case being misdiagnosed (*2*, *3*).

We used the generated data set to train an ensemble statistical model to predict selection coefficients based on the 6 parameters described. 25% of the simulated data sets were held back as an out-of-sample data set to be used for evaluating the performance of the trained statistical models and to test for overfitting. The remaining 75% of the simulated data was used for training three different statistical models (shape constrained additive models (SCAM), bagged multivariate regression splines (MARS), and Bayesian regularised neural networks (BRNN)) to predict selection coefficients using the six varied transmission model parameters. Statistical model performance was evaluated based on the root mean-squared error (RMSE). Optimum model-fitting hyperparameters based on RMSE were first identified by scanning over hyperparameters for each model before fitting each model. When identifying hyperparameters and training the final model, K-fold cross-validation sets were produced by splitting the training data into 20 sets of training data with the results of the cross-validation subsequently averaged to reduce any bias from the cross-validation set chosen. We calculated the performance of each trained model by calculating the RMSE for each model when tested using the holdout data set. To construct our final ensemble model, we simply calculated the average across the three models, weighted by their RMSE from the holdout test.

Uncertainty in selection coefficients due to stochastic variation in model simulations was also estimated using a similar statistical modelling framework (75% data split, hyperparameter tuning and 20-fold cross validation). For each parameter set, we used each trained model to first predict the selection coefficient. Next, we calculate the absolute prediction error by comparing the model prediction against the selection coefficient for each stochastic realisation, before calculating the standard deviation in the error across stochastic realisations. We trained a Bayesian regularised neural network model to predict the standard deviation in error before calculating robust confidence intervals given by ±1.96 * standard deviation.

To estimate the global risk of *pfhrp2/3* deletions spreading globally, we used the weighted average ensemble model to predict selection coefficients for each admin level 1 region based on their malaria prevalence and treatment related data, as detailed in the literature review and data sources section. A complete schematic of this modelling pipeline is given in Supplementary Figure 15.

### Supplementary Appendix 2. Database of model parameters associated with the strength of selection for pfhrp2/3 gene deletions

#### Introduction

This review aims to provide a database of relevant risk factors and disease epidemiology relevant for modelling the risk posed by the selection of pfhrp2/3 deletions. The review aims to identify, where available, estimates at the first administrative unit for key risk factors. If estimates are not available at the first administrative unit, national level estimates will be sourced. Estimates that are unavailable are recorded as missing.

The secondary aim of this review is to identify the sources of data available with which to estimate each parameter and to assess whether these provide additional data or insight beyond those that have been collected and modelled by the Malaria Atlas Project (MAP) (*1*) as part of their Commodities forecast modelling exercise (*2*).The MAP commodities database provides estimates of the treatment cascade at a national level, largely parameterised using Demographic and Health Surveys (DHS), Malaria Indicator Surveys (MIS), DHIS2 and World Malaria Report (WMR) data alongside a range of admin-level socio-economic covariates sourced from the Institute for Health Metrics and Evaluation used for fitting statistical models to these data. The specific treatment cascade estimates produced relevant for the modelling of pfhrp2/3 deletions of:

1. The probability that an individual will choose to seek care for fever. This decision probability is agnostic to the cause of fever.
2. The probability that an individual seeking care will seek treatment at a public vs private clinic.
3. The probability that an individual presenting at a private/public clinic will receive a diagnostic test.
4. The probability that the diagnostic test used is an RDT.
5. The probability that a tested individual will receive antimalarial treatment.

6. The probability that an untested individual will receive antimalarial treatment.

#### Methodology

In response, for the scope of this review, we consider the following risk factors. A high level overview of the data sources that we identified as part of our review for each factor are also provided:

| Number | Factor | Data sources Identified |
|---|---|---|
| #1 | Proportion of microscopy versus hrp2-based rapid diagnostic test (RDT) | WHO Global Health Observatory (GHO)<br><br>Literature review |
| #2 | Proportion of hrp2-based RDTs among all RDTs used | The Global Fund's Price and Quality Reporting (PQR) and President's Malaria Initiative Volume Data. |
| #3 | Proportion of children with fever who sought treatment advice | Demographic Health Surveys (DHS) |
| #4 | Proportion of suspected malaria cases who received any diagnostic test | WHO GHO |
| #5 | Malaria prevalence in children 2-10 years old | Malaria Atlas Project (MAP) |
| #6 | Proportion of RDT-negative suspected cases that did not receive any antimalarials (adherence to RDT results) | MAP<br><br>Literature review |
| #7 | Antimalarial market share of private versus public sector | ACTWatch project<br><br>Literature review |

#### Description of search strategy and selection criteria

The review was conducted initially with a scoping review of suitable databases already available for each of the factors, which have been collated by international organisations and entities such as WHO, The Global Fund, IHME and DHS. After identifying databases where available, we proceeded in the following two parts. Firstly, we retrieved and standardised data from databases identified in the scoping review at the subnational level or national level where appropriate. Secondly, we searched for estimates from the literature to fill in gaps (either at the subnational or national level where required) or to update data with more recent estimates than those identified in the second step. Given the availability of malaria prevalence and commodity estimates available from MAP for 2020, we aimed to collect estimates across all factors for 2020 where possible.

### Part 1 - Data from trusted sources

**Factor #1, #4:** Diagnostic testing and current usage of microscopy versus RDT

To assess the current usage of light microscopy and RDTs in malaria endemic countries, we used data from WHO Global Health Observatory (*3*) as of 02 February 2023 as well as literature published from 2021 to 2023. The following data streams were retrieved:

- Number of suspected malaria cases (#suspects)
- Number of suspected malaria cases who were examined by microscopy (#microscopy)
- Number of suspected malaria cases who were examined by RDTs (#rdt)

To calculate the testing coverage (factor #4) and the proportion of microscopy usage among all tests (factor #1), the following formulas were used:

$$testing\ coverage = \frac{\#microsscopy + \#rdt}{\#suspects}$$

$$proportion\ of\ microscopy = \frac{\#microscopy}{\#microscopy + \#rdt}$$

**Factor #2:** Distribution of RDT brands

To assess the market of RDTs in a country, data was obtained from The Global Fund's Price and Quality Reports (*4*) and data shared from the President's Malaria Initiative on RDT volumes distributed to each country. Records without any specific destination countries or

RDT brand names were removed. The target antigens of each RDT product were entered according to their reports on WHO Public Reports for In Vitro Diagnostics database (*3*). The proportions of all RDTs used in the last three years of data that were HRP2-based RDTs was estimated for each country.

**Factor #3:** Treatment seeking rate

It is important to assess treatment seeking behaviour of the population at risk since this allows for estimation of the effective coverage of RDTs as well as antimalarials. Data for this factor was extracted from the Demographic and Health Surveys (DHS) (*3*). Mixed records of national and subnational level were compiled. The indicator of interest is the percentage of under 5 febrile cases who sought any advice or treatment.

**Factor #5:** Malaria prevalence

To assess transmission intensity of a region, estimates for prevalence of *P.falciparum* malaria in children from 2 to 10 years old were obtained from Malaria Atlas Project (MAP) (*5*).

**Factor #6:** Level of adherence to RDT-negative results

Estimates for the proportion of RDT-negative cases who do not get any antimalarials were retrieved from MAP's Case Management Cascade[1].

**Factor #7:** Private-public market share of antimalarials

Information on public and private antimalarial market share was obtained from publications from the ACTwatch project[2]. An additional literature review was conducted for studies published from 2015 to 2023.

---

[1] https://data.malariaatlas.org/case-management
[2] https://www.actwatch.org/publications

#### Part 2 - Data from recent published literature

Several categories were used to construct search queries for publications on PubMed.

| Category | PubMed search terms |
|---|---|
| Publication from year X to year Y | (X:Y[pdat]) |
| Malaria | ("Malaria"[Mesh] OR malaria*[tiab] OR Plasmodium*[tiab] OR "malaria, falciparum"[MeSH Terms]) |
| Fever | ("Fever/diagnosis"[Mesh] OR "Fever/epidemiology"[Mesh] OR "Fever/therapy"[Mesh]) |
| Diagnostic test | ("diagnostic tests, routine"[MeSH Terms] OR "Malaria/diagnosis"[MAJR] OR ("diagnostic"[All Fields] AND "tests"[All Fields] AND "routine"[All Fields]) OR "routine diagnostic tests"[All Fields] OR ("diagnostic"[All Fields] AND "test"[All Fields]) OR "diagnostic test"[All Fields]) |
| Microscopy | ("microscopies"[All Fields] OR "microscopy"[MeSH Terms] OR "microscopy"[All Fields]) |
| RDT brand | (("First Response"[All Fields] OR "CareStart"[All Fields] OR "CareStart Malaria"[All Fields] OR "Bioline"[All Fields] OR "Bioline Malaria"[All Fields] OR "SD Bioline"[All Fields] OR "Paracheck"[All Fields] OR "Paracheck Pf"[All Fields] OR "ParaHIT"[All Fields] OR "ICT Malaria"[All Fields] OR “Advy”[All Fields] OR "STANDARD Q"[All Fields] OR "One Step"[All Fields] OR "Abbott"[All Fields] OR "NanoSign"[All Fields] OR "Malaria Ag"[All Fields] OR “Standard Diagnostics”[All Fields] OR "Onsite Pf/Pv"[All Fields] OR "FirstSign -"[All Fields] OR "OptiMAL-IT"[All Fields] OR "Clearview Malaria"[All Fields] OR "Malaria P.f/Pan"[All Fields] OR "SD BIOLINE"[All Fields] OR "Biocredit"[All Fields] OR "Hexagon Malaria"[All Fields] OR "ParaHIT- f"[All Fields] OR "Parahit f"[All Fields] OR "Parahit-f"[All Fields] OR "Onsite Pf"[All Fields] OR "Bionote"[All Fields] OR "ICT Diagnostics"[All Fields] OR "Paramax-3"[All Fields] OR "Parascreen"[All Fields] OR "Meriscreen"[All Fields] OR "Falcivax"[All Fields] OR "BinaxNOW Malaria"[All Fields] OR "Loopamp Malaria"[All Fields] OR "Humasis Malaria"[All Fields]) AND "rapid"[All Fields]) |
| Treatment | ("Malaria/drug therapy"[Mesh] OR "Antimalarials"[Mesh] OR "artemisinins"[MeSH Terms]) |
| Private sector | ("private sector"[MeSH Terms] OR ("private"[All Fields] AND "sector"[All Fields]) OR "private sector"[All Fields] OR "pharmacies"[MeSH Terms] OR pharmacy[All Fields] OR pharmacies[All Fields]) |
| Public sector | ("public sector"[MeSH Terms] OR ("public"[All Fields] AND "sector"[All Fields]) OR "public sector"[All Fields]) |
| Pfhrp2/3 deletion | ("pfhrp*"[All Fields] AND ("deletable"[All Fields] OR "deletant"[All Fields] |

| | |
|---|---|
| | OR "deletants"[All Fields] OR "delete"[All Fields] OR "deleted"[All Fields] OR "deleter"[All Fields] OR "deleters"[All Fields] OR "deletes"[All Fields] OR "deleting"[All Fields] OR "deletion s"[All Fields] OR "deletional"[All Fields] OR "gene deletion"[MeSH Terms] OR ("gene"[All Fields] AND "deletion"[All Fields]) OR "gene deletion"[All Fields] OR "deletions"[All Fields] OR "sequence deletion"[MeSH Terms] OR ("sequence"[All Fields] AND "deletion"[All Fields]) OR "sequence deletion"[All Fields] OR "deletion"[All Fields])) |
| Treatment seeking behaviour | ("patient acceptance of health care"[MeSH Terms] OR ("patient"[All Fields] AND "acceptance"[All Fields] AND "health"[All Fields] AND "care"[All Fields]) OR "patient acceptance of health care"[All Fields] OR ("treatment"[All Fields] AND "seeking"[All Fields] AND "behavior"[All Fields]) OR ("treatment"[All Fields] AND "seeking"[All Fields] AND "behaviour"[All Fields]) OR "treatment seeking behavior"[All Fields] OR "treatment seeking behaviour"[All Fields]) |
| Case management | ("case management"[MeSH Terms] OR ("case"[All Fields] AND "management"[All Fields]) OR "case management"[All Fields] OR "population surveillance"[MeSH Terms] OR ("therapy"[Subheading] OR "disease management"[MeSH Terms])) |
| Adherence | ("Guideline Adherence"[Mesh] OR "adhere"[All Fields] OR "adhered"[All Fields] OR "adherence"[All Fields] OR "adherences"[All Fields] OR "adherent"[All Fields] OR "adherents"[All Fields] OR "adherer"[All Fields] OR "adherers"[All Fields] OR "adheres"[All Fields] OR "adhering"[All Fields] OR "guideline adherence"[MeSH]) |
| Compliance | ("compliances"[All Fields] OR "patient compliance"[MeSH Terms] OR ("patient"[All Fields] AND "compliance"[All Fields]) OR "patient compliance"[All Fields] OR "compliance"[All Fields] OR "compliance"[MeSH Terms]) |
| Health knowledge and practice | ("health knowledge, attitudes, practice"[MeSH Terms]) |

Below are the inclusion and exclusion criteria for publications where additional literature reviews were done:

| Factor | Inclusion criteria | Exclusion criteria |
|---|---|---|
| 1 | ● Report the number of samples screened by either RDTs or microscopy<br>● Report on public sector<br>● Published from 2021 to 2023<br>● Are in English | ● Do not report the number of samples screened by either RDTs or microscopy<br>● Do not report on public sector<br>● Published before 2021<br>● Are not in English |
| 2 | ● Report the RDT brand used to diagnose P.falciparum infection<br>● Conduct in the following countries: Afghanistan, Angola, Burkina Faso, Bangladesh, Cote d'Ivoire, Cameroon, Congo, Democratic Republic of Congo, Comoros, Cabo Verde, Ethiopia, Ghana, Gambia, Guinea-Bissau, Guatemala, Guyana, Honduras, Haiti, Indonesia, Laos, Mali, Mozambique, Malawi, Niger, Nigeria, Nicaragua, Nepal, Pakistan, Philippines, Papua New Guinea, Democratic People's Republic of Korea, Zanzibar, Senegal, Solomon Islands, Sierra Leone, Somalia, South Sudan, Sao Tome and Principe, Eswatini, Chad, Togo, Tanzania, Zimbabwe, Burundi, Benin, Bolivia, Bhutan, Djibouti, Eritrea, Guinea, Kenya, Madagascar, Myanmar, Mauritania, Sudan, Timor-Leste, Uganda, Venezuela, Zambia<br>● Report sample collection year<br>● Published from 2020 to 2023<br>● Are in English | ● Do not report the RDT brand used to diagnose P.falciparum infection<br>● Do not conduct in the following countries: Afghanistan, Angola, Burkina Faso, Bangladesh, Cote d'Ivoire, Cameroon, Congo, Democratic Republic of Congo, Comoros, Cabo Verde, Ethiopia, Ghana, Gambia, Guinea-Bissau, Guatemala, Guyana, Honduras, Haiti, Indonesia, Laos, Mali, Mozambique, Malawi, Niger, Nigeria, Nicaragua, Nepal, Pakistan, Philippines, Papua New Guinea, Democratic People's Republic of Korea, Zanzibar, Senegal, Solomon Islands, Sierra Leone, Somalia, South Sudan, Sao Tome and Principe, Eswatini, Chad, Togo, Tanzania, Zimbabwe, Burundi, Benin, Bolivia, Bhutan, Djibouti, Eritrea, Guinea, Kenya, Madagascar, Myanmar, Mauritania, Sudan, Timor-Leste, Uganda, Venezuela, Zambia<br>● Do not report sample collection year<br>● Published before 2020<br>● Are not in English |
| 3 | ● Report the number/percentage of febrile children who sought care at public facilities<br>● Are not part of DHS program<br>● Published from 2010 to 2023<br>● Are in English | ● Do not report the number/percentage of febrile children who sought care at public facilities<br>● Are part of DHS program<br>● Published before 2010<br>● Are not in English |
| 4 | ● Report the number of samples screened by either RDTs or microscopy<br>● Report on public sector<br>● Published from 2021 to 2023 | ● Do not report the number of samples screened by either RDTs or microscopy<br>● Do not report on public sector<br>● Published before 2021<br>● Are not in English |

| | | |
|---|---|---|
| | • Are in English | |
| 6 | • Report the number of cases tested with RDT<br>• Report the number of RDT negative<br>• Report the number of RDT-negative cases who were (not) given antimalarials<br>• Published from 2015 to 2023<br>• Are in English | • Do not report the number of cases tested with RDT<br>• Do not report the number of RDT negative<br>• Do not report the number of RDT-negative cases who were (not) given antimalarials<br>• Published from before 2015<br>• Are not in English |
| 7 | • Report the number private facilities<br>• Report the number public facilities<br>• Published from 2010 to 2023<br>• Are not part of ACTWatch Project<br>• Are in English | • Do not report the number private facilities<br>• Do not report the number public facilities<br>• Published before 2010<br>• Are part of ACTWatch Project<br>• Are not in English |

The following table shows the final search queries used to retrieve publications relating to each risk factor:

| Factor | Search structure | Total hits | Included |
|---|---|---|---|
| 1 | <publication from 2021 to 2023> AND <malaria> AND (<diagnostic test> OR <microscopy>) AND <public sector> | 16 | 4 |
| 2 | ("Afghanistan"[Mesh] OR "Angola"[Mesh] OR "Burkina Faso"[Mesh] OR "Bangladesh"[Mesh] OR "Cote d'Ivoire"[Mesh] OR "Cameroon"[Mesh] OR "Congo"[Mesh] OR "Democratic Republic of the Congo"[Mesh] OR "Comoros"[Mesh] OR "Cabo Verde"[Mesh] OR "Ethiopia"[Mesh] OR "Ghana"[Mesh] OR "Gambia"[Mesh] OR "Guinea-Bissau"[Mesh] OR "Guatemala"[Mesh] OR "Guyana"[Mesh] OR "Honduras"[Mesh] OR "Haiti"[Mesh] OR "Indonesia"[Mesh] OR "Laos"[Mesh] OR "Mali"[Mesh] OR "Mozambique"[Mesh] OR "Malawi"[Mesh] OR "Niger"[Mesh] OR "Nigeria"[Mesh] OR "Nicaragua"[Mesh] OR "Nepal"[Mesh] OR "Pakistan"[Mesh] OR "Philippines"[Mesh] OR "Papua New Guinea"[Mesh] OR "Democratic People's Republic of Korea"[Mesh] OR "Zanzibar"[All Fields] OR "Senegal"[Mesh] OR "Melanesia"[MeSH Terms] OR “Solomon Islands”[All Fields] OR "Sierra Leone"[Mesh] OR "Somalia"[Mesh] OR "South Sudan"[Mesh] OR "Sao Tome and Principe"[Mesh] OR "Eswatini"[Mesh] OR “Swaziland”[All Fields] OR "Chad"[Mesh] OR "Togo"[Mesh] OR "Tanzania"[Mesh] OR "Zimbabwe"[Mesh] OR “Burundi"[Mesh] OR "Benin"[Mesh] OR "Bolivia"[Mesh] OR "Bhutan"[Mesh] OR "Djibouti"[Mesh] OR "Eritrea"[Mesh] OR "Guinea"[Mesh] OR "Kenya"[Mesh] OR "Madagascar"[Mesh] OR "Myanmar"[Mesh] OR "Mauritania"[Mesh] OR "Sudan"[Mesh] OR "Timor-Leste"[Mesh] OR "Uganda"[Mesh] OR "Venezuela"[Mesh] OR "Zambia”[Mesh]) AND <publication from 2020 to 2023> AND <malaria> AND <diagnostic test> AND <RDT brand> | 31 | 25 |
| 3 | <publication from 2010 to 2023> AND (<malaria> OR <fever>) AND ("patient acceptance of health care"[MeSH Terms] OR ("patient"[All Fields] AND "acceptance"[All Fields] AND "health"[All Fields] AND "care"[All Fields]) OR "patient acceptance of health care"[All Fields] OR ("treatment"[All Fields] AND "seeking"[All Fields] AND "behavior"[All Fields]) OR ("treatment"[All Fields] AND "seeking"[All Fields] AND "behaviour"[All Fields]) OR "treatment seeking behavior"[All Fields] OR "treatment seeking behaviour"[All Fields]) AND <public sector> | 63 | 0 |

| | | | |
|---|---|---|---|
| 4 | <publication from 2021 to 2023> AND <malaria> AND (<diagnostic test> OR <microscopy>) AND <public sector> | 16 | 4 |
| 6 | <publication from 2010 to 2023> AND <malaria> AND <diagnostic test> AND "rapid"[All Fields] AND “negative”[All Fields] AND <treatment> AND (<case management> OR <adherence> OR <compliance> OR <health knowledge and practice>) NOT ("glucosephosphate dehydrogenase"[MeSH]) NOT ("cost-benefit analysis"[MeSH]) | 158 | 0 |
| 7 | <publication from 2010 to 2023> AND <malaria> AND <public sector> AND <private sector> AND (<diagnostic test> OR <treatment>) | 111 | 48 |

All included studies as well as the identified target antigen(s) of all RDT listed in volume distribution data are available in the results of the literature review.

### Results

#### Literature Review Finding Overview

Firstly, the majority of databases that we identified for sourcing parameters are the same as those used by the Malaria Atlas Project (MAP) as part of their Commodities forecast modelling exercise. These include reliance on DHS/MIS/MICS/AIS data for treatment seeking data and the WHO WMR/GHO for estimates on diagnostic testing and RDT vs microscopy use.

With regards to adherence to RDT diagnostic test outcome, the only suitable data source was that produced by MAP. From our literature review, we did not identify any studies since 2015 that passed inclusion criteria. Of the studies that did include relevant data but were prior to 2015, 10 studies were identified, with an average adherence to RDT test outcomes of 81% across studies. The only other study of mention, was the analysis of ACT consortium data between 2007-2012 (thus excluded) across 10 studies that showed between 27% - 100% adherence across studies, with an overall mean across studies of 74%.

RDT brand volume data provided by PMI and PQR reporting data provided RDT volume proportions for all countries in Africa with malaria except for Equatorial Guinea and Gabon. While 25 additional studies passed inclusion criteria for the literature search regarding RDT brands, none of the studies included data on Equatorial Guinea and Gabon. In addition, the included studies provided reports on the brands of RDTs used as part of specific scientific investigations and do not necessarily reflect national RDT types used.

Lastly, the literature search resulted in 48 includes studies regarding the size of the private vs public market with regards to RDTs, antimalarials and treatment seeking behaviour more broadly. Studies from 21 countries were found, of which 4 were from Asia. A range of different measures of the private vs public sector were observed, e.g. % RDT sales according to manufacturers that went to private vs public, the number of treatment facilities classified as private vs public, surveys of individuals regarding where they sought treatment, analysis of DHS Service Provision Assessment surveys. In addition, DHS data also provides reports on where treatment seeking was sought in a number of DHS survey rounds, which is the underlying data used by MAP when modelling test adherence.

#### Decisions made regarding parameters for modelling exercise

Based on the findings from the literature review, we made the following decisions regarding how we parameterised each of the factors identified, as detailed in the table below.

| Number | Factor | Data sources used |
|---|---|---|
| #1 | Proportion of microscopy versus hrp2-based rapid diagnostic test (RDT) used | MAP commodities database |
| #2 | Proportion of hrp2-based RDTs among all RDTs used. | The Global Fund's Price and Quality Reporting (PQR) and President's Malaria Initiative Volume Data. |
| #3 | Proportion of children with fever who sought treatment advice | MAP commodities database and DHS data |
| #4 | Proportion of suspected malaria cases who received any diagnostic test | MAP commodities database |
| #5 | Malaria prevalence in children 2-10 years old | Malaria Atlas Project |
| #6 | Proportion of RDT-negative suspected cases that did not receive any antimalarials (adherence to RDT results) | MAP commodities database |
| #7 | Antimalarial market share of private versus public sector | MAP commodities database |

In overview, we made the decision to use the estimates provided by the MAP commodities database where available, with the notable exception being the use of Global Fund PQR and PMI RDT volume data, which was not reported by MAP. For factors #1, #3 and #4, our literature review identified the same underlying data sources. For factor #6, no studies passed inclusion criteria, and those that failed but had relevant data showed low non-adherence to RDT test outcomes, similar to the findings from the MAP Commodity forecasting database. Lastly, for factor #7, we did identify a number of studies, however, the range of different metrics reported and incomplete coverage across countries led to us making the decision to use the MAP commodity estimates here. This has the advantage of using estimates that are consistent across the different outputs, as the methodology used by MAP is self consistent. Additionally, the results of our literature review indicate that ultimately a statistical model is required to provide estimates for each country, as needed for our exercise. Consequently, we have determined that the database reflects our best understanding at a global level of the factors known to be important for the selection of *pfhrp2/3* deletions.

For disclaimer purposes, this decision was reached independently by the first three authors of the paper, before contacting individuals from Malaria Atlas Project for further guidance on interpretation of commodity estimates and uncertainty estimates.

### Final Pipeline for sourcing estimates for the hrp2/3 modelling exercise

As inputs for the malaria transmission model, we require the following parameters for each level one administrative unit:

1. Malaria prevalence
2. Effective Treatment Seeking - defined as the probability that an individual with symptomatic malaria, seeks treatment and is treated.
3. The use of microscopy for diagnostic testing
4. Nonadherence to diagnostic test outcomes
5. Proportion of RDT brands used that only target PfHRP2 (Prospective Risk Score only)

**Malaria prevalence**

We use the median and 95% confidence interval of slide prevalence 2-10 years old for 2020 estimated by MAP (*5*).

**Effective Treatment Seeking**

We do not explicitly model the private and public markets for treatment and consequently we aggregate over both sources for treatment to estimate an overall estimate. We use MAP commodity dashboard estimates for each country at both the private and public level, with the overall effective treatment seeking given by:

$$f_T = (f_{S\_Private} * T_{Private} * Tr_{PosTest\_Private}) +$$
$$(f_{S\_Private} * (1-T_{Private}) * Tr_{UnTest\_Private}) +$$
$$(f_{S\_Public} * (T_{Public}) * Tr_{Pos\_Public}) +$$
$$(f_{S\_Public} * (1-T_{Public}) * Tr_{UnTest\_Publ})$$

Where $f_T$ is the national effective treatment seeking probability, $f_{S\_Private}$ is care-seeking in private sector, $f_{S\_Public}$ is care-seeking in public sector, $T_{Private}$ is testing probability in private sector, $T_{Public}$ is testing probability in public sector, $Tr_{PosTest\_Private}$ is the treatment probability given a positive test in the private sector, $Tr_{PosTest\_Public}$ is the treatment probability given a positive test in the public sector, $Tr_{UnTest\_Private}$ is the probability of treatment in the private sector given an individual was not tested and $Tr_{UnTest\_Public}$ is the probability of treatment in the public sector.

$f_T$ calculated in this way provides a national estimate, however, DHS surveys provide subnational data on care-seeking overall and are shown to exhibit large ranges in some countries. To address, this we build a gradient boosted tree model to predict the DHS care-seeking probability in under 5s ($f_S$) in the most recent DHS surveys available using maps of healthcare travel time and friction surface maps previously estimated by MAP (*6*), population weighted to first administrative unit level using World Population Project estimates (*7*), and a subset of national level covariates collected by the Economist as part of their excess mortality modelling exercise (*8*), which includes covariates on healthcare spending, wealth, democracy, urbanisation, inequality and other socio-economic indicators of healthcare access and capacity. We use 20-fold cross validation, trained on a sunset of 75% of the data, with tuned hyperparameters: a learning rate of 0.05, max tree depth of 12, subsample ratio of the training instance data of 0.85, and subsample ratio of columns when constructing each tree of 0.85.

We use the trained model to predict subnational estimates of care-seeking probabilities in under 5s, $f_S$ (i.e. the DHS metric), in countries without DHS surveys. In order to ensure the national, population weighted estimate of $f_T$ is consistent with the MAP commodity estimates, the value of $f_{T\_i}$ in administrative unit *i* (where there are *n* administrative units in total) is subsequently given by:

$$f_{S_i} * \frac{f_T * pop_{national}}{\sum_{i=1}^{n} f_{S_i} * pop_i}$$

In this way, we are able to provide subnational estimates of effective treatment seeking that reflect both DHS subnational patterns and the statistical modelling framework used as part of the MAP commodity forecasts exercise.

**Microscopy use for diagnosis**

The MAP commodity dashboard provides estimates of the proportion of diagnostic tests conducted using RDTs ($RDT_{prop}$). Given the noted difficulties in the use of diagnostic testing in the private sector (*9*), we make the simplifying assumption that the majority of microscopy use is concentrated in the public sector. Consequently, we model the overall proportion of microscopy use as:

$$\frac{(1-RDT_{prop})^{*}f_{S\_Public}}{f_{S\_Public}+f_{S\_Private}}$$

**Nonadherence to diagnostic test outcomes**

The MAP commodity dashboard provides modelled estimates of the proportion of individuals who are treated despite producing a negative diagnostic test in both the private, $Tr_{PosTest\_Private}$, and public $Tr_{PosTest\_Public}$ sector. We use these values to model the overall probability of a negative RDT still being treated, i.e. non adherence to the diagnostic test.

$$\frac{(Tr_{NegTestTreat\_Public}{}^{*}f_{S\_Public}}{f_{S\_Public}+f_{S\_Private}}+\frac{(Tr_{NegTestTreat\_Private}{}^{*}f_{S\_Private}}{f_{S\_Public}+f_{S\_Private}}$$

**Proportion of RDT brands used that only target PfHRP2 (Prospective Risk Score only)**

Lastly, to estimate the proportion of HRP2-based RDTs (brands that target only PfHRP2), we estimated the proportion of RDT volumes (sourced from The Global Fund's Price and Quality Reporting (PQR) and shared from the President's Malaria Initiative Volume Data) distributed to each country between 2018 - 2021 that target only PfHRP2.

**Uncertainty Intervals**

We accounted for uncertainty in model parameters as follows. For treatment seeking, MAP provided for each country the relative change in all cause (private and public) care seeking estimates directly from their statistical model fitting from the median reflected by the 95% credible intervals. Similar estimates were provided for their estimates of microscopy use and test non-adherence. Both were used to construct upper and lower bounds for effective treatment seeking and microscopy use in each country. For malaria prevalence, we used the 95% confidence intervals of slide prevalence 2-10 years old provided publicly by MAP (*5*). No uncertainty was available for proportions of RDT brands used that only target PfHRP2 and consequently we did not include this in our prospective modelling. Lastly, we estimated 95% credible intervals for fitness costs and HRP3 cross reactivity as part of our model fitting exercise to surveillance data in Ethiopia and Eritrea.

3. Malaria testing and diagnosis, (available at https://www.who.int/data/gho/data/themes/topics/topic-details/GHO/malaria-testing-diagnosis).

4. Workbook: Price & quality reporting transaction summary, (available at https://insights.theglobalfund.org/t/Public/views/PriceQualityReportingTransactionSummary/TransactionSummary?iframeSizedToWindow=true&:embed=y&:showAppBanner=false&:display_count=no&:showVizHome=no).

5. Malaria Atlas Project Pf Infection Prevalence (per 100 Children). *Malaria Atlas Project*, (available at https://data.malariaatlas.org/trends?year=2020&metricGroup=Malaria&geographicLevel=admin1&metricSubcategory=Pf&metricType=rate&metricName=PR).

6. D. J. Weiss, A. Nelson, H. S. Gibson, W. Temperley, S. Peedell, A. Lieber, M. Hancher, E. Poyart, S. Belchior, N. Fullman, B. Mappin, U. Dalrymple, J. Rozier, T. C. D. Lucas, R. E. Howes, L. S. Tusting, S. Y. Kang, E. Cameron, D. Bisanzio, K. E. Battle, S. Bhatt, P. W. Gething, A global map of travel time to cities to assess inequalities in accessibility in 2015. *Nature*. **553**, 333–336 (2018).

7. World Population Prospects - Population Division - United Nations, (available at https://population.un.org/wpp/).

8. T. Economist, S. U. Solstad, The pandemic’s true death toll. [online] The Economist. Available at: https://www.economist.com/graphic-detail/coronavirus-excess-deaths-estimates [Accessed 12 June 2023]. First published in the article “Counting the dead”, The Economist, issue 20, 2021. *Economist* (available at https://github.com/TheEconomist/covid-19-the-economist-global-excess-deaths-model).

9. Meeting report of the WHO technical consultation on malaria case management in the private sector in high-burden countries (1-3 May 2019), (available at https://www.who.int/publications/m/item/WHO-CDS-GMP-MPAC-2019.12).
